# SARS-CoV-2 vaccination in the first year after hematopoietic cell transplant or chimeric antigen receptor T cell therapy: A prospective, multicenter, observational study (BMT CTN 2101)

**DOI:** 10.1101/2024.01.24.24301058

**Authors:** Joshua A. Hill, Michael J. Martens, Jo-Anne H. Young, Kavita Bhavsar, Jianqun Kou, Min Chen, Lik Wee Lee, Aliyah Baluch, Madhav V. Dhodapkar, Ryotaro Nakamura, Kristin Peyton, Dianna S. Howard, Uroosa Ibrahim, Zainab Shahid, Paul Armistead, Peter Westervelt, John McCarty, Joseph McGuirk, Mehdi Hamadani, Susan DeWolf, Kinga Hosszu, Elad Sharon, Ashley Spahn, Amir A. Toor, Stephanie Waldvogel, Lee M. Greenberger, Jeffery J. Auletta, Mary M. Horowitz, Marcie L. Riches, Miguel-Angel Perales

## Abstract

**Background:** The optimal timing of vaccination with SARS-CoV-2 vaccines after cellular therapy is incompletely understood.

**Objective:** To describe humoral and cellular responses after SARS-CoV-2 vaccination initiated <4 months versus 4-12 months after cellular therapy.

**Design:** Multicenter prospective observational study.

**Setting:** 34 centers in the United States.

**Participants:** 466 allogeneic hematopoietic cell transplant (HCT; n=231), autologous HCT (n=170), or chimeric antigen receptor T cell (CAR-T cell) therapy (n=65) recipients enrolled between April 2021 and June 2022.

**Interventions:** SARS-CoV-2 vaccination as part of routine care.

**Measurements:** We obtained blood prior to and after vaccinations at up to five time points and tested for SARS-CoV-2 spike (anti-S) IgG in all participants and neutralizing antibodies for Wuhan D614G, Delta B.1.617.2, and Omicron B.1.1.529 strains, as well as SARS-CoV-2-specific T cell receptors (TCRs), in a subgroup.

**Results:** Anti-S IgG and neutralizing antibody responses increased with vaccination in HCT recipients irrespective of vaccine initiation timing but were unchanged in CAR-T cell recipients initiating vaccines within 4 months. Anti-S IgG ≥2,500 U/mL was correlated with high neutralizing antibody titers and attained by the last time point in 70%, 69%, and 34% of allogeneic HCT, autologous HCT, and CAR-T cell recipients, respectively. SARS-CoV-2-specific T cell responses were attained in 57%, 83%, and 58%, respectively. Humoral and cellular responses did not significantly differ among participants initiating vaccinations <4 months vs 4-12 months after cellular therapy. Pre-cellular therapy SARS-CoV-2 infection or vaccination were key predictors of post-cellular therapy anti-S IgG levels.

**Limitations:** The majority of participants were adults and received mRNA vaccines.

**Conclusions:** These data support starting mRNA SARS-CoV-2 vaccination three to four months after allogeneic HCT, autologous HCT, and CAR-T cell therapy.

**Funding:** National Marrow Donor Program, Leukemia and Lymphoma Society, Multiple Myeloma Research Foundation, Novartis, LabCorp, American Society for Transplantation and Cellular Therapy, Adaptive Biotechnologies, and the National Institutes of Health

## INTRODUCTION

Recipients of cellular therapies, including hematopoietic cell transplant (HCT) and chimeric antigen receptor T cells (CAR-T cells), have a high risk for morbidity from infection with severe acute respiratory syndrome coronavirus 2 (SARS-CoV-2), the cause of coronavirus disease 2019 (Covid-19) (1–4). Without pre-existing immunity from previous infection or vaccination, up to 30% of cellular therapy recipients die within 4-6 weeks after infection with SARS-CoV-2 (5,6).

Initial studies of SARS-CoV-2 vaccination demonstrated low immunogenicity in patients with hematologic malignancies receiving chemotherapy and cellular therapies (7–12). Most studies focused on humoral immunity using binding antibody measurements, but evaluation of neutralizing antibodies and T cell responses provide additional data relevant to protection from severe infection (13,14). Studies in cellular therapy recipients have few participants vaccinated within the first 3-12 months, precluding conclusions about the optimal timing of vaccination after cellular therapy (15–28).

Although immune responses to routine vaccines following HCT are known to be diminished, guidelines recommend inactivated vaccines such as influenza as soon as three months after HCT and other inactivated vaccines as early as six months post-HCT (29–33). Therefore, guidelines recommended SARS-CoV-2 re-vaccination as early as three months post-cellular therapy in the absence of real-world data (34,35). While knowledge is rapidly accumulating (36–39), the optimal timing, schedule of vaccination, and immunological correlates for protective immunity after cellular therapy are unknown. To address these knowledge gaps, the Center for International Blood and Marrow Transplant Research (CIBMTR) and Blood and Marrow Transplant Clinical Trials Network (BMT CTN) conducted a multi-center, prospective, observational study of the safety and immunogenicity of SARS-CoV-2 vaccination within 12 months after autologous HCT, allogeneic HCT, and CAR-T cell therapy. We previously reported the findings among the first 175 allogeneic HCT recipients (40). In this report, we present the final analyses from the complete cohort of allogeneic HCT recipients in addition to autologous HCT and CAR-T cell recipients.

## METHODS

### Participants and study design

We prospectively enrolled patients of any age who underwent an allogeneic HCT, autologous HCT, or CAR-T cell therapy and were planning to receive a first post-cellular therapy SARS-CoV-2 vaccine within 12 months of treatment. Type of SARS-CoV-2 vaccine, number of doses, and timing post-HCT were at the discretion of institutional standards or provider preference. The study (CIBMTR SC21-07/BMT CTN 2101) was approved by the Institutional Review Board of the National Marrow Donor Program and opened to enrollment in April 2021. Participants or their legal guardians provided written informed consent. This study follows the Strengthening the Reporting of Observational Studies in Epidemiology (STROBE) reporting guideline for observational studies.

### Procedures

Blood was collected within pre-specified windows of two weeks prior to first vaccination (pre-V1), at least 3 weeks after first vaccination and within one week prior to second vaccination (post-V1), one to five weeks after second and third vaccination (post-V2 and post-V3, respectively), and seven to nine months after enrollment (end-of-study) (**Figure S1**). Data collection is detailed in the Supplement.

### Testing

#### Binding and neutralizing antibodies

We tested serum for semiquantitative total IgG to the SARS-CoV-2 spike protein (S) receptor-binding domain with the Roche Elecsys Anti-SARS-CoV-2 S assay (anti-S IgG), qualitative detection of high-affinity antibodies to SARS-CoV-2 nucleocapsid (N) protein (anti-N IgG), and neutralizing antibodies for Wuhan D614G, Delta B.1.617.2, and Omicron B.1.1.529 strains (LabCorp; Burlington, NC; Supplement) (41). The upper limit of quantitation for anti-S IgG was 2,500 U/mL. All samples were tested for anti-S IgG; only baseline samples were tested for anti-N IgG. Neutralizing antibodies were tested at the pre-V1, post-V2, and post-V3 or end-of-study time points in up to 30 chronologically enrolled participants per cellular therapy cohort and vaccine initiation timing subgroup.

#### SARS-CoV-2-specific T cells

We performed T cell receptor (TCR) variable beta chain immunosequencing of genomic DNA from peripheral blood mononuclear cells (PBMCs) using the ImmunoSEQ Assay (Adaptive Biotechnologies, Seattle, WA) to quantify the absolute abundance of unique SARS-CoV-2–specific TCRs as previously described (42–44). We quantified SARS-CoV-2 TCR breadth, defined as the proportion of total unique TCRs associated with SARS-CoV-2, and depth, defined as the extent to which SARS-CoV-2-associated TCRs expand. Samples were classified as positive, negative, or “no call” (representing samples with insufficient TCR rearrangements) using the T-Detect classifier (Adaptive Biotechnologies, Seattle, WA) based on breadth and depth compared to a reference population of individuals with prior SARS-CoV-2 infection. Testing was performed in the same subgroup of individuals tested for neutralizing antibodies.

#### Multiparametric Flow Cytometric Analysis

Cryopreserved PBMCs from pre-V1 samples were tested by flow cytometry to quantitate CD19+ B cells and CD4+ T cells (Supplement).

### Statistical analysis

The primary objective was to compare the immunogenicity of SARS-CoV-2 vaccines in patients starting <4 months versus 4-12 months after cellular therapy. Power calculations based on anticipated and actual sample sizes are in the Supplement and **Tables S1-S2**.

To determine relevant anti-S IgG thresholds for immunogenicity, receiver operating characteristic (ROC) curves were employed using anti-S IgG as a continuous marker and dichotomous outcomes of neutralizing antibodies at the median level (≥5,274 ID50) achieved in a non-immunocompromised cohort vaccinated with two doses of mRNA-1273 (Moderna) in a clinical trial using the same assay (41). The proportions of participants with anti-S IgG levels above this threshold (defined as a positive response) are described with Wald 99% confidence intervals (CIs); response rates were compared using a two-sided Z test of the difference in proportions between timing strata. To adjust for imbalances in baseline variables between timing cohorts, propensity scores for the likelihood of being in the <4-month cohort were constructed using logistic regression. A propensity-adjusted analysis compared positive responses at the post-V2, post-V3, and end-of-study time points between the <4-month and 4–12-month subgroups within each cellular therapy cohort. For each timing subgroup, inverse probability weights were constructed from the reciprocal of the propensity of being included in the cohort; weighted response proportions and their standard errors were computed and used to obtain point estimates and 99% CIs for the propensity-adjusted difference in response rates. We used adjusted logistic and linear regression models to evaluate the impact of vaccination timing and cellular therapy type on anti-S IgG and T cell responses. Results from samples obtained within six months of administration of the monoclonal antibody tixagevimab-cilgavimab (Evusheld, AstraZeneca) were excluded from antibody analyses. P-values <0.01 were used to determine statistical significance to account for multiple comparisons. Analyses were performed using SAS Version 9.4 and R version 4.2.

## RESULTS

### Participants and treatment characteristics

We enrolled 231 allogeneic HCT, 170 autologous HCT, and 65 CAR-T cell recipients (N=466 in total) who received ≥1 SARS-CoV-2 vaccine at 34 centers in the United States between April 2021 and June 2022. Overall, 231 (50%) participants were vaccinated <4 months after cellular therapy and 235 (50%) 4–12 months after cellular therapy. Demographic and baseline characteristics are in **Table 1** and were similar within each cohort stratified by vaccination <4 months versus 4-12 months after cellular therapy (data not shown). Allogeneic HCT recipients were enrolled earlier in the pandemic than autologous HCT and CAR-T cell recipients. Most allogeneic HCT recipients (77%) were taking immunosuppressive medications at baseline. Characteristics of the subgroup of 151 participants (60 allogeneic HCT, 53 autologous HCT, and 38 CAR-T cell recipients) tested for neutralizing antibodies and SARS-CoV-2-specific T cells were similar to the overall cohort (data not shown).

**Table 1.** Demographic and clinical characteristics of the participants.

| **Characteristic** | **Allogeneic HCT** | **Autologous HCT** | **CAR-T** | **Total** |
| --- | --- | --- | --- | --- |
| No. of patients | 231 | 170 | 65 | 466 |
| Age at cellular therapy, years - no. (%) |  |  |  |  |
| Median (range) | 56.5 (3.0-76.7) | 60.6 (7.0-77.9) | 62.7 (11.0-82.8) | 58.6 (3.0-82.8) |
| <18 | 20 (9) | 3 (2) | 2 (3) | 25 (5) |
| 18-29 | 20 (9) | 9 (5) | 1 (2) | 30 (6) |
| 30-39 | 23 (10) | 9 (5) | 2 (3) | 34 (7) |
| 40-49 | 26 (11) | 18 (11) | 7 (11) | 51 (11) |
| 50-59 | 50 (22) | 43 (25) | 19 (29) | 112 (24) |
| 60-69 | 71 (31) | 63 (37) | 17 (26) | 151 (32) |
| ≥70 | 21 (9) | 25 (15) | 17 (26) | 63 (14) |
| Sex - no. (%) |  |  | 1.00 |  |
| Male | 120 (52) | 97 (57) | 35 (54) | 252 (54) |
| Female | 111 (48) | 73 (43) | 30 (46) | 214 (46) |
| Hispanic or Latinx ethnicity- no. (%) |  |  |  |  |
| Yes | 14 (6) | 13 (8) | 5 (8) | 32 (7) |
| No | 216 (94) | 157 (92) | 60 (92) | 433 (93) |
| Unknown/Not reported | 1 (0) | 0 (0) | 0 (0) | 1 (0) |
| Race or Ethnic Group other than Hispanic or Latinx - no. (%) |  |  |  |  |
| White | 175 (76) | 124 (73) | 53 (82) | 352 (76) |
| Black | 30 (13) | 33 (19) | 7 (11) | 70 (15) |
| Asian | 15 (6) | 6 (4) | 4 (6) | 25 (5) |
| American Indian or Alaskan Native | 2 (1) | 1 (1) | 1 (2) | 4 (1) |
| More than one race | 3 (1) | 1 (1) | 0 (0) | 4 (1) |
| Unknown/Not reported | 6 (3) | 5 (3) | 0 (0) | 11 (2) |
| Underlying Disease - no. (%) |  |  |  |  |
| AML/MDS/MPN | 147 (64) | 0 (0) | 0 (0) | 147 (32) |
| ALL/other leukemia/aplastic anemia | 59 (26) | 0 (0) | 5 (8) | 64 (14) |
| Non-Hodgkin lymphoma | 14 (6) | 32 (19) | 37 (57) | 81 (17) |
| Multiple myeloma | 0 (0) | 117 (69) | 23 (35) | 140 (30) |
| Others | 11 (5) | 21 (12) | 0 (0) | 32 (7) |
| Date of cellular therapy - no. (%) |  |  |  |  |
| May, 2020 - December, 2020 | 22 (10) | 6 (4) | 2 (3) | 30 (6) |
| January, 2021 - March, 2021 | 53 (23) | 15 (9) | 3 (5) | 71 (15) |
| April, 2021 - June, 2021 | 80 (35) | 42 (25) | 9 (14) | 131 (28) |
| July, 2021 - September, 2021 | 56 (24) | 35 (21) | 11 (17) | 102 (22) |
| October, 2021 – December, 2021 | 12 (5) | 56 (33) | 19 (29) | 87 (19) |
| January, 2022 – June, 2022 | 8 (3) | 16 (9) | 21 (32) | 45 (10) |
| Number of enrolling centers | 30 | 28 | 18 | 34 |
| Graft source - no. (%) |  |  |  |  |
| Bone marrow | 37 (16) | 1 (1) | 0 (0) | 38 (8) |
| Peripheral blood | 185 (80) | 169 (99) | 65 (100) | 419 (90) |

| **Characteristic** | **Allogeneic HCT** | **Autologous HCT** | **CAR-T** | **Total** |
| --- | --- | --- | --- | --- |
| Cord blood | 9 (4) | 0 (0) | 0 (0) | 9 (2) |
| Donor and HLA match - no. (%) |  |  |  |  |
| Matched related | 42 (18) | N/A | N/A | N/A |
| Matched unrelated | 107 (46) | N/A | N/A | N/A |
| Mismatched related | 63 (27) | N/A | N/A | N/A |
| Mismatched unrelated | 19 (8) | N/A | N/A | N/A |
| Conditioning intensity - no. (%) |  |  |  |  |
| Myeloablative | 109 (47) | N/A | N/A | N/A |
| Reduced intensity | 81 (35) | N/A | N/A | N/A |
| Non-myeloablative or none | 41 (18) | N/A | N/A | N/A |
| GVHD prophylaxis - no. (%) |  |  |  |  |
| Post-HCT cyclophosphamide regimen | 90 (39) | N/A | N/A | N/A |
| Non-post-HCT cyclophosphamide regimen | 129 (56) | N/A | N/A | N/A |
| T cell depletion | 12 (5) | N/A | N/A | N/A |
| Acute GVHD prior to baseline sample - no. (%) |  |  |  |  |
| No | 144 (62) | N/A | N/A | N/A |
| Yes | 87 (38) | N/A | N/A | N/A |
| Chronic GVHD prior to baseline sample - no. (%) |  |  |  |  |
| No | 206 (89) | N/A | N/A | N/A |
| Yes | 25 (11) | N/A | N/A | N/A |
| Immunosuppressive medications in use - no. (%) |  |  |  |  |
| Pre-V1 | 179 (77) | 47 (28) | 7 (11) | 233 (50) |
| Post-V2 | 162 (70) | 67 (39) | 6 (9) | 235 (50) |
| Post-V3 | 179 (77) | 89 (52) | 9 (14) | 277 (59) |
| Post-V4 | 92 (40) | 70 (41) | 5 (8) | 167 (63) |
| End-of-study | 96 (42) | 75 (44) | 10 (15) | 181 (39) |
| Absolute lymphocyte count (cells/mm<sup>3</sup>) at baseline, median (IQR) | 700 (440-1110) | 1100 (700-1640) | 605 (320-900) | 800 (500-1280) |
| Absolute CD19+ B cell count (cells/mm<sup>3</sup>) at baseline, median (IQR) | 89 (41-183) | 139 (46-276) | 12 (3-50) | 84 (34-198) |
| Absolute CD4+ T cell count (cells/mm<sup>3</sup>) at baseline, median (IQR) | 170 (64-404) | 514 (245-870) | 194 (90-516) | 271 (108-624) |

Percentages may not total 100 because of rounding. HCT indicates hematopoietic cell transplant; IQR, interquartile range; AML, acute myelogenous leukemia; MDS, myelodysplastic syndrome; MPN, myeloproliferative neoplasm; ALL, acute lymphoblastic leukemia; HLA, human leukocyte antigen; GVHD, graft-versus-host disease.

SARS-CoV-2 vaccination, infection, and treatment characteristics of study participants are in **Table 2**. The distribution of vaccination timing is depicted in **Figure S2.** Vaccination for SARS-CoV-2 prior to receipt of cellular therapy was more frequent in the <4-month subgroups, consistent with the observation that these participants tended to be enrolled at a later calendar time when SARS-CoV-2 vaccines were more available. Most participants (n=356, 76%) received ≥3 vaccine doses, and the distribution of the number of vaccine doses was similar in those vaccinated <4 months versus 4-12 months after cellular therapy. At baseline, anti-N IgG was positive in 100 (21%) individuals, least frequent in allogeneic HCT recipients, but similar across vaccine initiation time subgroups. Tixagevimab-cilgavimab and intravenous immunoglobulin (IVIG) were administered prior to one or more samples in 29% and 14% of participants, respectively; both were more common in the CAR-T cell cohort but similar across vaccine initiation time subgroups. SARS-CoV-2 infections were identified in 99 participants after initiating vaccinations and were equally distributed across vaccine timing subgroups but more frequent after CAR-T cell therapy (**Table 2**).

**Table 2.**
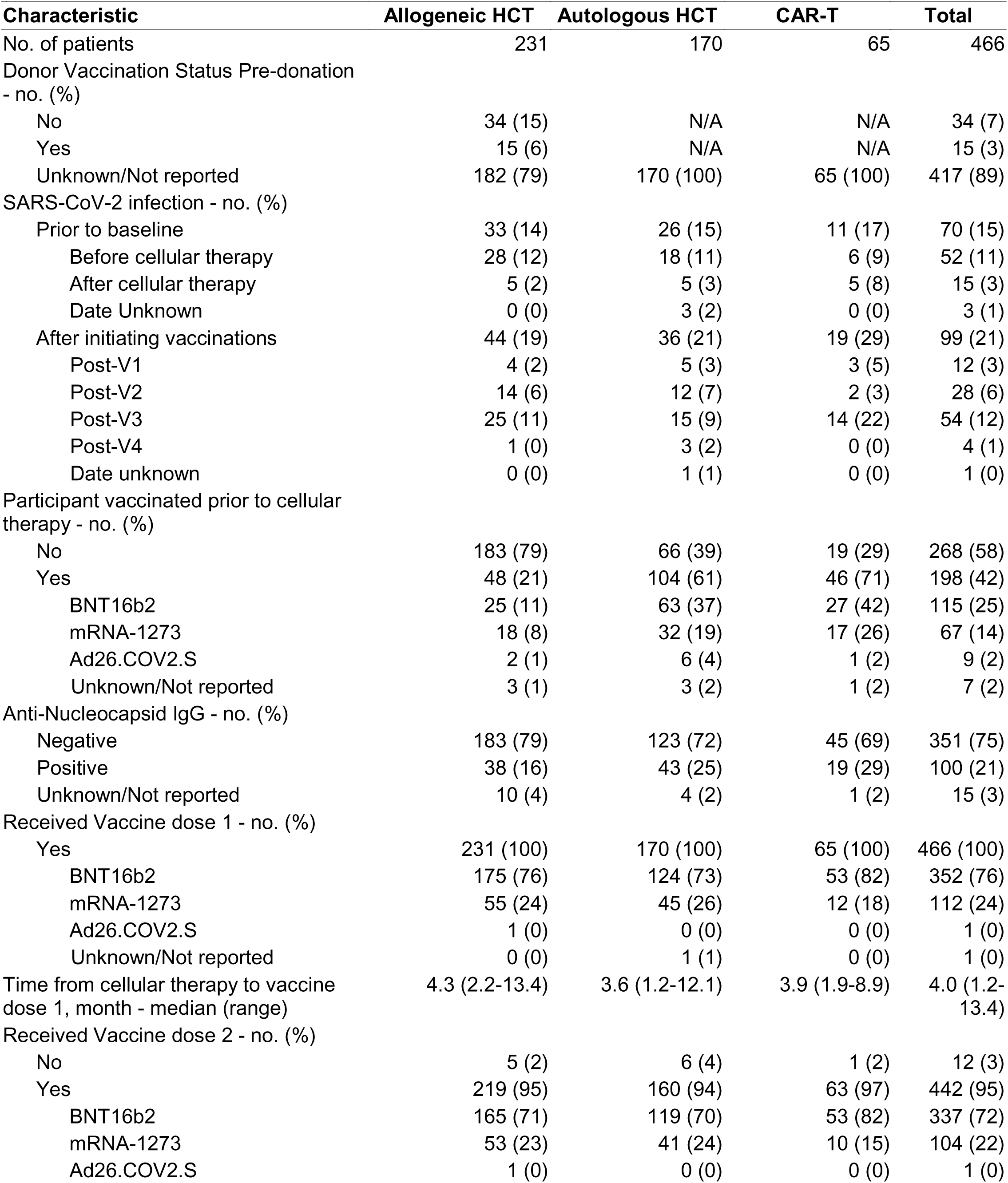
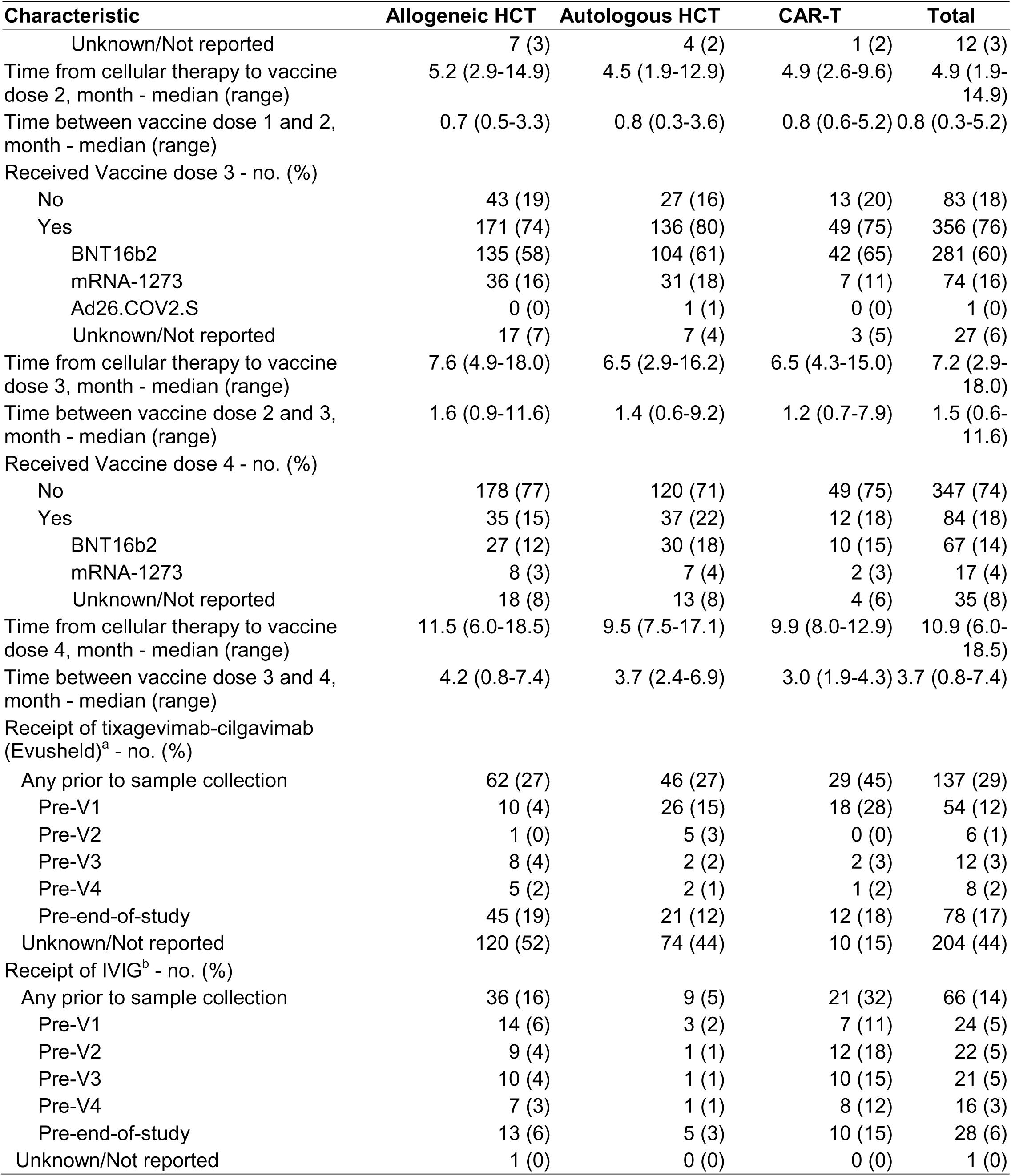
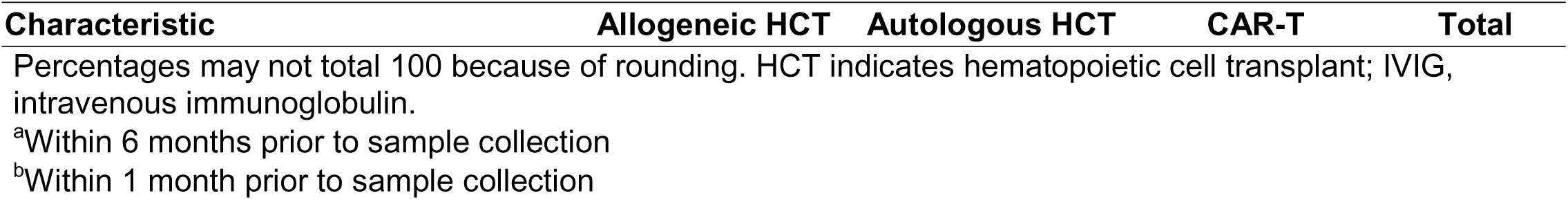
SARS-CoV-2 vaccination, infection, and treatment characteristics of the participants.

### Binding and neutralizing antibody responses increased with subsequent vaccination in HCT recipients irrespective of vaccine initiation timing but were unchanged in CAR-T cell recipients initiating vaccines within 4 months

Median anti-S IgG titers were higher in the <4-month subgroups at the baseline (pre-V1) time point in all cohorts but only significantly different in allogeneic HCT recipients (**Figure 1A**). This difference persisted at the post-V1 time point, but subsequent measurements were similar in all cohorts among those starting vaccines <4 months versus 4-12 months after cellular therapy (**Table S3)**. Anti-S IgG increased at post-V2 and subsequent time points in both vaccine timing subgroups among the allogeneic and autologous HCT cohorts (**Table S4**). In the CAR-T cell cohort, anti-S IgG titers were relatively unchanged after vaccinations in the <4-month subgroup; in the 4-12-month subgroup, titers increased after two vaccines with a subsequent plateau, but the changes did not reach statistical significance. Results in the CAR-T cell cohort stratified by CD19 versus BCMA-targeted therapies are in **Figure S3**.

**Figure 1.**
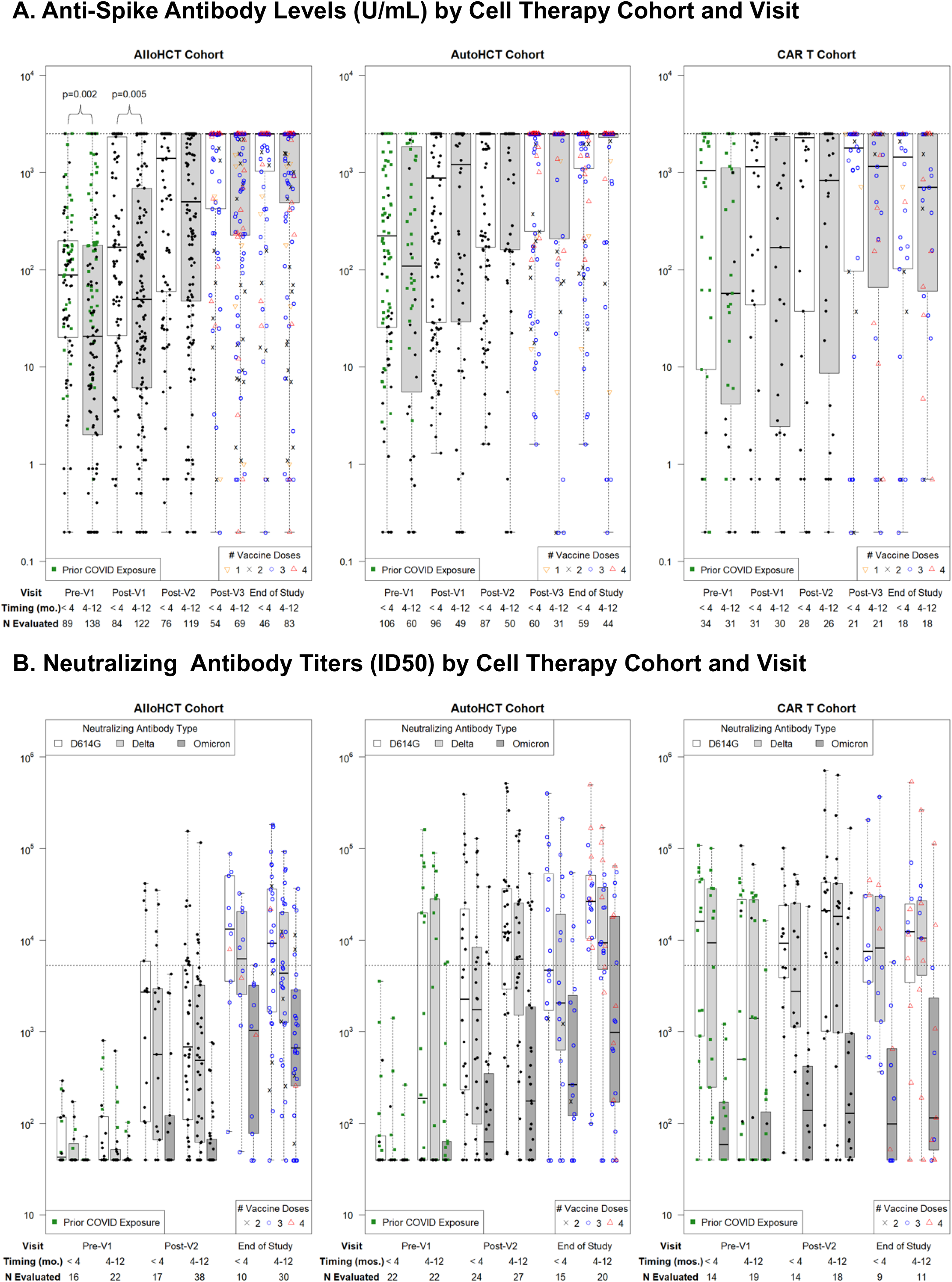
Longitudinal SARS-CoV-2 anti-Spike IgG titers and neutralizing antibody titers stratified by cellular therapy cohort and vaccine initiation timing. **A)** SARS-CoV-2 anti-S IgG titers per time point. The horizontal dotted line indicates the threshold for a positive response, defined as anti-S IgG >2,500 U/mL as determined from a ROC curve analysis; this was also the upper limit of quantitation for the assay. **B)** SARS-CoV-2 neutralizing antibody titers (ID50) in a subgroup of 151 participants; ID50 is defined as the reciprocal of the sample dilution required to reduce relative luminescence units by 50%. The horizontal dotted line shows the median neutralizing antibody level (5,274 ID50) achieved in a healthy cohort vaccinated with two doses of mRNA-1273 (Moderna) in a clinical trial and tested with the same assay and defined here as a positive response. In panels A and B, ‘prior COVID exposure’ (green squares) indicates data in the first two time points from participants with a known prior SARS-CoV-2 infection, prior SARS-Cov-2 vaccination in the participant or hematopoietic cell donor, or positive anti-N IgG assay at baseline. Results are depicted on a log_10_ scale. Time points tested within 6 months of receipt of tixagevimab-cilgavimab (Evusheld) were excluded.

Neutralizing antibody titers increased with vaccinations in the allogeneic and autologous HCT cohorts and were similar at the end-of-study time point by vaccine initiation timing (**Figure 1B; Table S3-S4**). In the CAR-T cell cohort, similar patterns were seen as for the anti-S IgG results and were comparable at the end-of-study time point by vaccine initiation timing. In all cohorts, most participants had low neutralizing antibody levels for Omicron B.1.1.529.

### The proportion of participants with positive antibody responses increased with vaccination and was similar at the end-of-study time point after HCT, irrespective of vaccine initiation timing; CAR-T cell recipients had declining proportions despite vaccination

An ROC curve analysis demonstrated that anti-S IgG performed well for predicting high-level neutralizing antibodies with area under the curve (AUC) values of 0.93 and 0.89 in the allogeneic and autologous HCT cohorts, respectively, and 0.72 in the CAR-T cell cohort (**Figure S4)**. Based on these analyses, we defined an anti-S IgG threshold of >2,500 U/mL as a positive response due to correlation with development of high-level neutralizing antibodies across all cohorts.

A positive response was attained in 70%, 69%, and 34% of allogeneic HCT, autologous HCT, and CAR-T cell recipients by the last time point in propensity-adjusted analyses that accounted for imbalances in baseline variables between vaccine initiation timing subgroups (**Figure 2**). Response rates were similar when stratified by vaccine initiation <4 months versus 4-12 months after allogeneic and autologous HCT. In the CAR-T cell group, response rates were twice as high in the <4-month subgroup at the last time point; however, similar differences were seen at the baseline time point, and the proportion of CAR-T cell recipients with anti-S IgG ≥2,500 U/mL declined over time in both subgroups despite vaccination.

**Figure 2.**
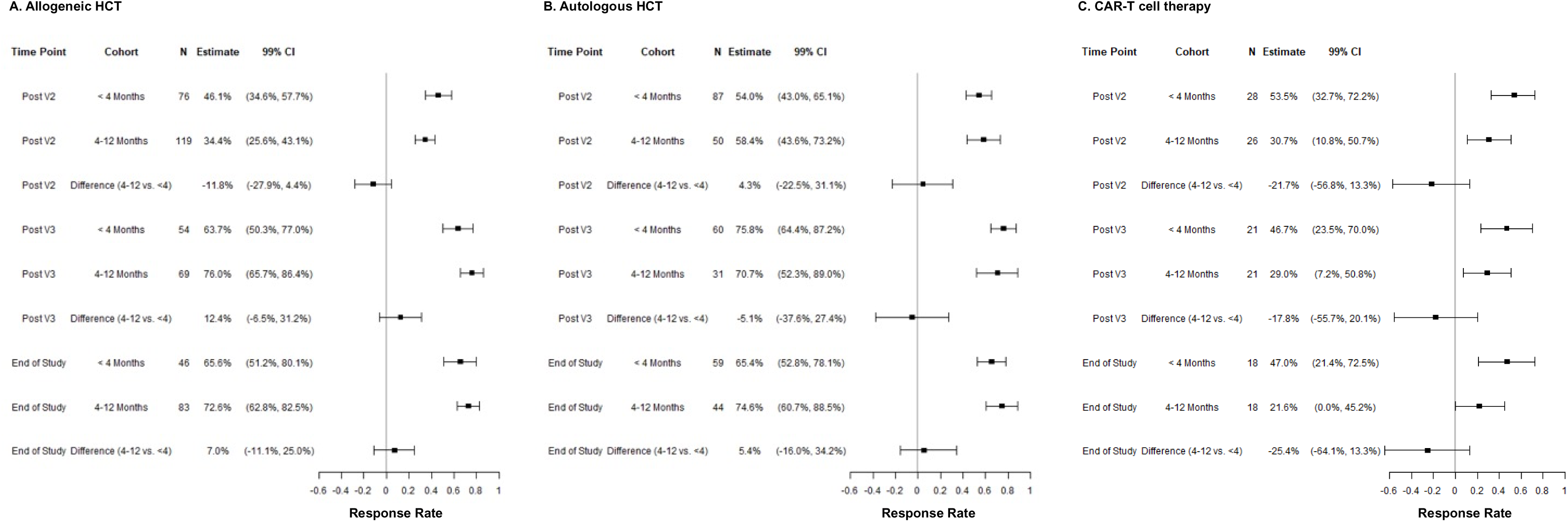
Propensity score-adjusted antibody response rates (anti-Spike IgG >2,500 U/mL) after two or more vaccine doses stratified by cellular therapy cohort and vaccine initiation timing. Forest plots of the proportion of individuals at each time point, stratified by vaccine initiation <4 months versus 4-12 months after cellular therapy, who had a positive anti-Spike IgG value (defined as >2,500 U/mL). The allogeneic HCT, autologous HCT, and CAR-T cell therapy cohorts are depicted in Panels **A**, **B**, and **C**, respectively. Propensity scores of being in the <4-month timing subgroup were constructed using stepwise variable selection with a criterion of a p-value ≤0.05 to determine which variables were included in the final model. Potential interactions were evaluated between covariates. Cell therapy type, lymphocyte count, and calendar time of enrollment were included in the final model. Inverse probability weights (IPW) were constructed from these propensity scores for all patients according to their timing groups. These were used to obtain adjusted response rate estimates and confidence intervals using IPW proportions and their standard errors. Wald 99% confidence intervals (CI) are shown. Comparisons used a Wald test to compare IPW weighted proportions. Patients who received tixagevimab-cilgavimab (Evusheld) within the prior six months were excluded.

### SARS-CoV-2-specific T cell responses increased with vaccination in all cohorts and were similar by vaccine initiation timing

In the subgroup of 151 participants (60 allogeneic HCT, 53 autologous HCT, and 38 CAR-T cell recipients) tested for SARS-CoV-2-specific TCRs, the pre-V1 sample was positive by the T-Detect test assay in three of 60 (5%) allogeneic HCT recipients, 12 of 53 (23%) autologous HCT recipients, and 11 of 38 (29%) CAR-T cell recipients (**Figure 3A**). This is consistent with the observation that autologous HCT and CAR-T cell recipients were enrolled later compared to allogeneic HCT recipients with more frequent prior vaccination or infection.

**Figure 3.**
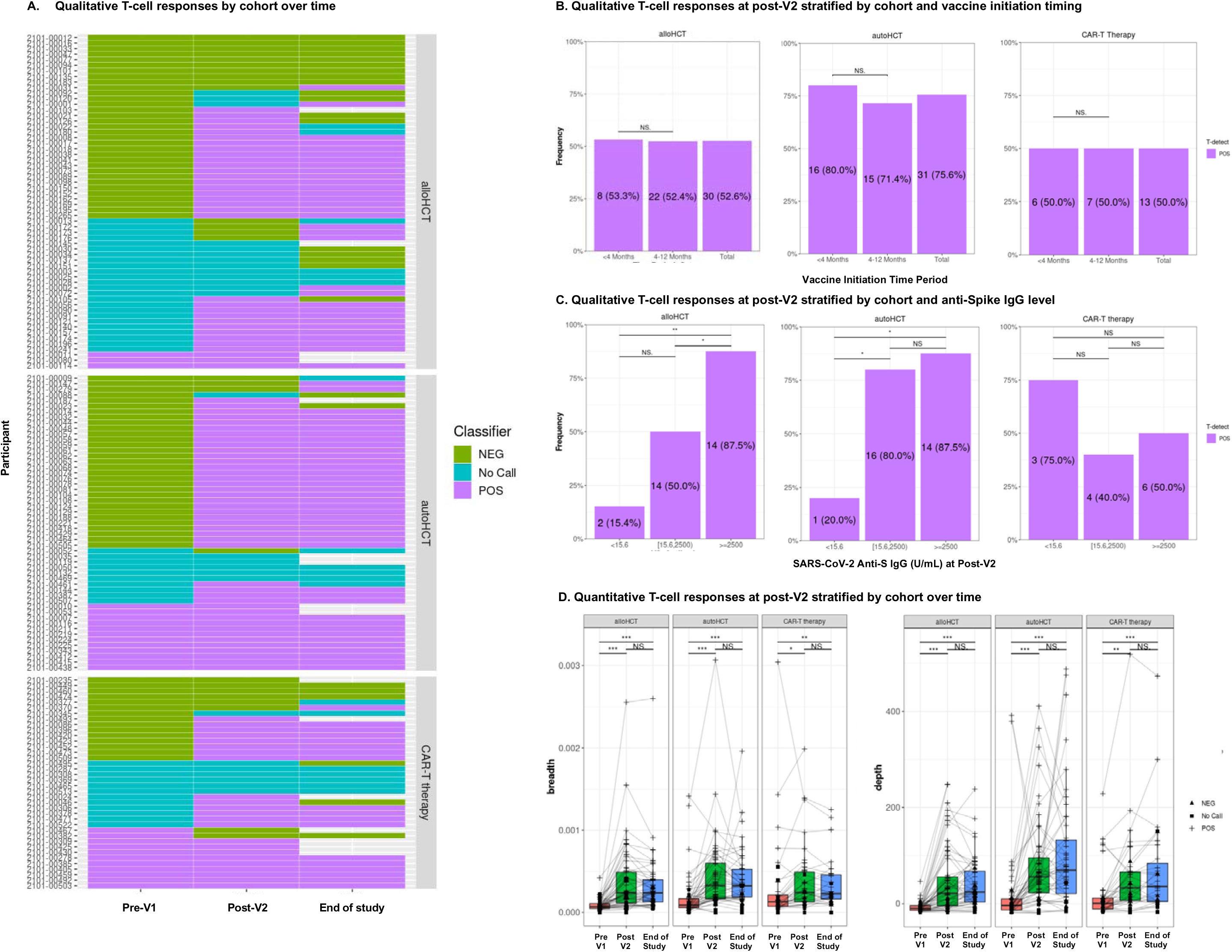
SARS-CoV-2-specific T cell receptor (TCR) variable beta chain sequencing results in a subgroup of 151 participants. **A)** Qualitative results indicating a positive, negative, or indeterminate (‘no call’) result for the presence of SARS-CoV-2-specific TCRs based on the T-Detect ImmunoSEQ Assay classifier (Adaptive Biotechnologies, Seattle, WA). Each row indicates a unique participant clustered by allogeneic HCT recipients at the top of the panel, autologous HCT recipients in the middle, and CAR-T cell therapy recipients at the bottom of the panel. Unfilled cells at the end-of-study time point indicate that no sample was available for testing. **B)** The proportion of participants with a positive T-Detect at the post-V2 timepoint stratified by cellular therapy cohort and vaccine initiation timing subgroup. Participants with a positive test at baseline were excluded from these analyses. **C**) The proportion of participants with a positive T-Detect at the post-V2 timepoint in categories of negative, any detectable, or positive (>2,500 U/mL) SARS-CoV-2 anti-S IgG titers. Any detectable antibody was based on a threshold determined to be predictive of detection of neutralizing antibodies at any level. In panels B and C, individuals with a positive T-Detect at the pre-V1 time point were excluded. (**D)** Quantitative values at each time point indicating the SARS-CoV-2 TCR breadth, defined as the proportion of total unique TCRs associated with SARS-CoV-2, and depth, defined as the extent to which SARS-CoV-2-associated TCRs expand. NS indicates not significant; *, ≤0.05; **, ≤0.01; ***, ≤0.001.

At the post-V2 time point, 55%, 81%, and 61% of allogeneic HCT, autologous HCT, and CAR-T cell recipients had a positive T-Detect, respectively, and findings were relatively unchanged at the end-of-study time point (**Table S5**). Similar proportions of HCT recipients had a positive T-Detect in the <4-month and 4-12-month subgroups; comparisons in the CAR-T cell cohort were limited by sample size (**Figure 3B**). The proportion of participants with a positive T-Detect increased when compared across categories of no, low, and positive SARS-CoV-2 anti-S IgG levels in the HCT cohorts, but a low proportion (15%-20%) had a positive T-Detect in the absence of antibodies (**Figure 3C**). Conversely, 75% of CAR-T cell recipients had a positive T-Detect in the absence of SARS-CoV-2 anti-S IgG, and there was no correlation between T cell responses and antibody titers, although observations were limited by sample size.

We next quantified the relative number (breadth) and relative sum frequency (depth) of detectable SARS-CoV-2-specific TCRs. We observed increased depth and breadth at the post-V2 time point in all three cohorts, with stable results at the end-of-study time point (**Figure 3D**). When stratified by categories of SARS-CoV-2 anti-S IgG titers, breadth and depth increased as the anti-S IgG titer increased in the allogeneic HCT and autologous HCT recipients but not CAR-T cell recipients (**Figure S5**). Metrics were similar in CD19 versus BCMA-CAR-T cell recipients at the post-V2 time point (**Figure S6**).

### Pre-cellular therapy SARS-CoV-2 infection or vaccination in the recipient are key predictors of post-cellular therapy SARS-CoV-2 humoral and cellular immunity

We performed multivariable logistic and linear regression models evaluating cellular therapy type, vaccine initiation timing, and other covariates on vaccine response as measured by SARS-CoV-2 anti-S IgG, both as a categorical variable (positive response above versus below 2,500 U/mL) and continuous variable at post-V2, post-V3, and end-of-study time points (**Table S6-S7**). There were no significant differences in quantitative SARS-CoV-2 anti-S IgG values by vaccine initiation timing subgroup within any of the cellular therapy cohorts. Antibody titers were lower in the CAR-T cell cohort than the HCT cohorts, particularly at the last time point. Throughout most models and time points, prior vaccination or infection with SARS-CoV-2 in the recipient, as well as absolute CD19^+^ B cell counts at enrollment, were the most significant predictors of higher SARS-CoV-2 anti-S IgG values. Similarly, prior SARS-CoV-2 vaccination or infection were associated with higher SARS-CoV-2-specific T cell responses (**Table S8**).

### Anti-S IgG and T cell responses correlated with subsequent SARS-CoV-2 infection

Comparisons of antibody and T cell measurements at the post-V2 and end-of-study time points with subsequent SARS-CoV-2 infection demonstrated that participants with lower anti-S IgG titers and with a negative T-Detect assay were more likely to be diagnosed with SARS-CoV-2, but the differences did not reach statistical significance (**Table S9)**. Infections were likely underestimated.

### SARS-CoV-2 mRNA vaccination was safe

Possible vaccine-related grade 3 or higher adverse events were uncommon (**Table S10)**. New-onset acute or chronic GVHD were within the expected ranges for this patient population. No CAR-T cell recipients experienced recurrent immune-associated toxicities attributed to vaccination, such as cytokine release syndrome or immune effector cell-associated neurotoxicity syndrome.

## DISCUSSION

In this prospective, multicenter study of 466 allogeneic HCT, autologous HCT, and CAR-T cell recipients who initiated SARS-CoV-2 vaccination with mRNA vaccines within the first 12 months after cellular therapy, we demonstrate that humoral and cellular responses after two or more vaccinations were similar in participants initiating vaccination within <4 months versus 4-12 months. Binding and neutralizing antibody responses increased over time in the HCT cohorts but remained relatively unchanged in the CAR-T cell cohort. However, SARS-CoV-2 specific T cells increased in depth and breadth with vaccination in all cohorts. In adjusted models, pre-cellular therapy SARS-CoV-2 vaccination or infection of the recipient and baseline B cell count were key predictors of post-cellular therapy immunity.

We previously reported results from the first 175 allogeneic HCT recipients enrolled in this study, which demonstrated similar findings of comparable humoral and cellular responses after two or more SARS-CoV-2 mRNA vaccinations initiated <4 months versus 4-12 months after HCT (40). In this manuscript, we include an expanded cohort of allogeneic HCT recipients with increased diversity in race, ethnicity, and age (more pediatric participants) in addition to autologous HCT and CAR-T cell cohorts. The results of our prior study were replicated in the expanded allogeneic HCT cohort and the autologous HCT cohort. Notably, there was no evidence for a humoral immune response in the CAR-T cell cohort initiating mRNA SARS-CoV-2 vaccination <4 months after CAR-T cell infusion. Although binding and neutralizing antibody titers increased overall in the 4-12 month vaccine initiation subgroup, the changes were not significant, and baseline levels started at a lower level than in the <4 month subgroup. In both CAR-T cell subgroups, the proportion of participants with an anti-S IgG titer >2,500 U/mL decreased from the baseline to the end-of-study time point despite vaccination, with only 33% of participants achieving this threshold. It is possible that vaccination maintained antibody titers at a higher level than would otherwise be anticipated after B cell depleting CAR-T cell therapy.

Beyond humoral immune responses, pathogen-specific T cell responses are a critical aspect of assessing immunity to viral pathogens in immunocompromised individuals. T cell responses play a key role in controlling the severity of disease once infected (45,46). An important finding in this study was the development of SARS-CoV-2-specific T cell responses in the majority of participants in all three cellular therapy cohorts, and responses appeared to peak after two vaccine doses, similar to findings in related contexts (7). The proportion of participants with a positive T-Detect assay was similar across vaccine initiation timing subgroups and generally increased when compared across categories of no, low, and positive SARS-CoV-2 anti-S IgG levels in the HCT cohorts, although was not correlated with antibody responses in the CAR-T cell cohort. In a study of T cell responses to SARS-CoV-2 vaccination and infection in individuals lacking B cells due to treatment with rituximab or a primary immunodeficiency, the authors observed greater reactivity and proliferative capacity of SARS-CoV-2-specific T cells elicited by both infection and vaccination in the B cell deficient patients compared to healthy controls (45). Furthermore, they observed a 4.8-fold reduced odds of moderate or severe Covid-19 in vaccinated B cell deficient participants despite the absence of vaccine-induced antibodies. Together, these data underscore the importance of vaccinating CAR-T cell recipients to establish cellular immunity despite an expectation of poor humoral immune responses.

In adjusted models evaluating the impact of cellular therapy type and vaccine initiation timing, we demonstrated that prior vaccination or infection with SARS-CoV-2 in the recipient were the most consistently significant predictors of higher SARS-CoV-2 anti-S IgG and T cell values. The persistence of pre-cellular therapy SARS-CoV-2-specific humoral immunity was apparent based on the frequent presence of antibodies at the baseline time point in all cohorts, with generally higher antibody titers in patients tested earlier after cellular therapy. Notably, we did not observe associations with other clinical variables often considered in immunocompromised patients, such as the presence of GVHD, use of immunosuppressive therapies, or absolute lymphocyte counts (15–18,20–22,24–26).

This study is one of the largest prospective analyses of vaccination within the first year after cellular therapy and supports current guidelines for SARS-CoV-2 vaccination starting approximately three months after cell infusion (35,47). Most guidelines recommend vaccine initiation six to twelve months after cellular therapies, although some advocate initiation as soon as three months (29–31,33,48,49). Our data provide encouraging proof-of-concept for the utility of early vaccination targeting additional pathogens with mRNA vaccine platforms and underscore the importance of investigating multiple aspects of immunogenicity. The observation that neutralizing antibody titers remained low for SARS-CoV-2 variants after Wuhan D614G-targeted vaccination underscores the importance of booster vaccinations, continued utilization and development of prophylactic and therapeutic interventions, and other infection prevention strategies in this population.

A limitation is that this was an observational study, and we may not have fully accounted for confounding, although we used rigorous statistical methodology to account for observed differences. We are unable to account for variables impacting vaccine initiation timing based on center policies or patient-specific considerations. Because vaccine practices evolved over the course of the study, participants recruited earlier in the study were more likely to have initiated vaccination ≥4 months following cellular therapy and less likely to have had prior vaccination or infection. This may explain in part the lower baseline anti-S IgG titers observed in the 4-12-month cohorts; however, we accounted for this variation using propensity-adjusted analyses. We also note that in the allogeneic HCT cohort, data were limited or unavailable for donors in regard to prior SARS-CoV-2 vaccination, prior infection, and SARS-CoV-2 anti-S IgG, which could impact recipient immunity (50,51). Comparisons in the CAR-T cell cohort had lower power due to fewer participants. There was attrition during followup, which could affect our findings. Only two individuals received non-mRNA vaccines, so these results only apply to SARS-CoV-2 mRNA vaccines, and a minority of participants received a fourth vaccine dose. There was limited enrollment of pediatric patients.

In conclusion, these data support ensuring appropriate vaccination of patients prior to receiving cellular therapy in addition to starting mRNA SARS-CoV-2 vaccination within three to four months after allogeneic HCT, autologous HCT, or CAR-T cell therapy.

## Contributors

JAH, MLR, M-AP, MMH, and MJM designed the study; JAH, MLR, M-AP, MMH, MJM, LMG, JHY, and JA interpreted the results; MJM, MC, LWL, JK, SDW, JAH, JHY, and MLR analyzed the data and created the figures; JAH, JHY, KB, AB, RN, KP, AS, PA, PW, DSH, UI, JM, CB, JM, MH, SDW, M-AP collected data; JAH, JHY, MLR drafted the manuscript. JAH, MJM, and JK accessed and verified the underlying data. All authors contributed to the writing and revision of the manuscript and approved the final version.

## Data Availability

These data will be publicly available either on the CIBMTR website or in BioLINCC, the CIBMTR and BMT CTN platforms for public sharing of data.

## Acknowledgments

We are grateful for the support provided by the study teams at all participating centers, the Research Advisory Committee, study investigators, and the research participants. The authors also thank the Data Operations and IT groups in CIBMTR (on both the Medical College of Wisconsin and NMDP campus) for their assistance.

## Data sharing

These data will be publically available either on the CIBMTR website or in BioLINCC, the CIBMTR and BMT CTN platforms for public sharing of data.

## REFERENCES

1. Robilotti E V., Babady NE, Mead PA, Rolling T, Perez-Johnston R, Bernardes M, et al. Determinants of COVID-19 disease severity in patients with cancer. Nat Med. 2020 Aug 1;26(8):1218–23.

2. Kuderer NM, Choueiri TK, Shah DP, Shyr Y, Rubinstein SM, Rivera DR, et al. Clinical impact of COVID-19 on patients with cancer (CCC19): a cohort study. The Lancet. 2020 Jun 20;395(10241):1907–18.

3. Mato AR, Roeker LE, Lamanna N, Allan JN, Leslie L, Pagel JM, et al. Outcomes of COVID-19 in patients with CLL: a multicenter international experience. Blood. 2020 Sep 3;136(10):1134–43.

4. Mushtaq MU, Shahzad M, Chaudhary SG, Luder M, Ahmed N, Abdelhakim H, et al. Impact of SARS-CoV-2 in Hematopoietic Stem Cell Transplantation and Chimeric Antigen Receptor T Cell Therapy Recipients. Transplant Cell Ther. 2021 Sep 1;27(9):796.e1–796.e7.

5. Sharma A, Bhatt NS, St Martin A, Abid MB, Bloomquist J, Chemaly RF, et al. Clinical characteristics and outcomes of COVID-19 in haematopoietic stem-cell transplantation recipients: an observational cohort study. Lancet Haematol. 2021 Mar 1;8(3):e185–93.

6. Infante MS, Nemirovsky D, Devlin S, DeWolf S, Tamari R, Dahi PB, et al. Outcomes and Management of the SARS-CoV2 Omicron Variant in Recipients of Hematopoietic Cell Transplantation and Chimeric Antigen Receptor T Cell Therapy. Transplant Cell Ther [Internet]. 2023 Oct [cited 2023 Dec 10]; Available from: https://pubmed.ncbi.nlm.nih.gov/37806446/

7. Greenberger LM, Saltzman LA, Gruenbaum LM, Xu J, Reddy ST, Senefeld JW, et al. Anti-spike T-cell and Antibody Responses to SARS-CoV-2 mRNA Vaccines in Patients with Hematologic Malignancies. Blood Cancer Discov. 2022 Sep 27;OF1–9.

8. Addeo A, Shah PK, Bordry N, Hudson RD, Albracht B, Di Marco M, et al. Immunogenicity of SARS-CoV-2 messenger RNA vaccines in patients with cancer. Cancer Cell. 2021 Jun;39(8):1091–1098.e2.

9. Herishanu Y, Avivi I, Aharon A, Shefer G, Levi S, Bronstein Y, et al. Efficacy of the BNT162b2 mRNA COVID-19 vaccine in patients with chronic lymphocytic leukemia. Blood. 2021 Jun 10;137(23):3165–73.

10. Pleyer C, Ali MA, Cohen JI, Tian X, Soto S, Ahn IE, et al. Effect of bruton tyrosine kinase inhibitor on efficacy of adjuvanted recombinant hepatitis B and zoster vaccines. Blood. 2021 Jan 14;137(2):185–9.

11. Thakkar A, Gonzalez-Lugo JD, Goradia N, Gali R, Shapiro LC, Pradhan K, et al. Seroconversion rates following COVID-19 vaccination amongst patients with cancer. Cancer Cell. 2021;

12. Haggenburg S, Hofsink Q, Lissenberg-Witte BI, Broers AEC, Doesum JA van, Binnendijk RS van, et al. Antibody Response in Immunocompromised Patients With Hematologic Cancers Who Received a 3-Dose mRNA-1273 Vaccination Schedule for COVID-19. JAMA Oncol [Internet]. 2022 Oct 1 [cited 2023 Oct 5];8(10):1477–83. Available from: https://jamanetwork.com/journals/jamaoncology/fullarticle/2795176

13. Dispinseri S, Secchi M, Pirillo MF, Tolazzi M, Borghi M, Brigatti C, et al. Neutralizing antibody responses to SARS-CoV-2 in symptomatic COVID-19 is persistent and critical for survival. Nat Commun. 2021 May 11;12(1):1–12.

14. Barouch DH. Covid-19 Vaccines — Immunity, Variants, Boosters. New England Journal of Medicine. 2022 Sep 15;387(11):1011–20.

15. Einarsdottir S, Martner A, Nicklasson M, Wiktorin HG, Arabpour M, Törnell A, et al. Reduced immunogenicity of a third COVID-19 vaccination among recipients of allogeneic hematopoietic stem cell transplantation. Vol. 107, Haematologica. Ferrata Storti Foundation; 2022. p. 1479–82.

16. Dhakal B, Abedin S, Fenske T, Chhabra S, Ledeboer N, Hari P, et al. Response to SARS-CoV-2 vaccination in patients after hematopoietic cell transplantation and CAR T-cell therapy. Blood. 2021 Oct 7;138(14):1278–81.

17. Bergman P, Blennow O, Hansson L, Mielke S, Nowak P, Chen P, et al. Safety and efficacy of the mRNA BNT162b2 vaccine against SARS-CoV-2 in five groups of immunocompromised patients and healthy controls in a prospective open-label clinical trial. EBioMedicine. 2021 Dec 1;74:103705.

18. Redjoul R, Le Bouter A, Beckerich F, Fourati S, Maury S. Antibody response after second BNT162b2 dose in allogeneic HSCT recipients. The Lancet. 2021 Jul 24;398(10297):298–9.

19. Le Bourgeois A, Coste-Burel M, Guillaume T, Peterlin P, Garnier A, Béné MC, et al. Safety and Antibody Response after 1 and 2 Doses of BNT162b2 mRNA Vaccine in Recipients of Allogeneic Hematopoietic Stem Cell Transplant. JAMA Netw Open. 2021;4(9).

20. Ram R, Hagin D, Kikozashvilli N, Freund T, Amit O, Bar-On Y, et al. Safety and Immunogenicity of the BNT162b2 mRNA COVID-19 Vaccine in Patients after Allogeneic HCT or CD19-based CART therapy—A Single-Center Prospective Cohort Study. Transplant Cell Ther. 2021 Jun 30;27(9):788–94.

21. Huang A, Cicin-Sain C, Pasin C, Epp S, Audigé A, Müller NJ, et al. Antibody Response to SARS-CoV-2 Vaccination in Patients Following Allogeneic Hematopoietic Cell Transplantation. Transplantation and Cellular Therapy, Official Publication of the American Society for Transplantation and Cellular Therapy. 2022 Jan;0(0).

22. Tamari R, Politikos I, Knorr DA, Vardhana SA, Young JC, Marcello LT, et al. Predictors of Humoral Response to SARS-CoV-2 Vaccination after Hematopoietic Cell Transplantation and CAR T-cell Therapy. Blood Cancer Discov. 2021 Nov 1;2(6):577–85.

23. Ali H, Ngo D, Aribi A, Arslan S, Dadwal S, Marcucci G, et al. Safety and Tolerability of SARS-CoV2 Emergency-Use Authorized Vaccines for Allogeneic Hematopoietic Stem Cell Transplant Recipients. Transplant Cell Ther. 2021 Jul;27(11):938.e1–938.e6.

24. Kimura M, Ferreira VH, Kothari S, Pasic I, Mattsson JI, Kulasingam V, et al. Safety and Immunogenicity After a Three-Dose SARS-CoV-2 Vaccine Schedule in Allogeneic Stem Cell Transplant Recipients. Transplant Cell Ther. 2022 Oct 1;28(10):706.e1–706.e10.

25. Mori Y, Uchida N, Harada T, Katayama Y, Wake A, Iwasaki H, et al. Predictors of impaired antibody response after SARS-CoV-2 mRNA vaccination in hematopoietic cell transplant recipients: A Japanese multicenter observational study. Am J Hematol [Internet]. 2023 Jan 1 [cited 2024 Jan 7];98(1):102–11. Available from: https://pubmed.ncbi.nlm.nih.gov/36260658/

26. Barkhordar M, Chahardouli B, Biglari A, Ahmadvand M, Bahri T, Alaeddini F, et al. Three doses of a recombinant conjugated SARS-CoV-2 vaccine early after allogeneic hematopoietic stem cell transplantation: predicting indicators of a high serologic response-a prospective, single-arm study. Front Immunol [Internet]. 2023 [cited 2024 Jan 7];14. Available from: https://pubmed.ncbi.nlm.nih.gov/37153556/

27. Piñana JL, Martino R, Vazquez L, López-Corral L, Pérez A, Chorão P, et al. SARS-CoV-2-reactive antibody waning, booster effect and breakthrough SARS-CoV-2 infection in hematopoietic stem cell transplant and cell therapy recipients at one year after vaccination. Bone Marrow Transplant [Internet]. 2023 May 1 [cited 2024 Jan 7];58(5):567. Available from: /pmc/articles/PMC9974060/

28. Aleissa MM, Little JS, Davey S, Saucier A, Zhou G, Gonzalez-Bocco IH, et al. Severe Acute Respiratory Syndrome Coronavirus 2 Vaccine Immunogenicity among Chimeric Antigen Receptor T Cell Therapy Recipients. Transplant Cell Ther [Internet]. 2023 Jun 1 [cited 2024 Jan 7];29(6):398.e1–398.e5. Available from: https://pubmed.ncbi.nlm.nih.gov/36906276/

29. Cordonnier C, Einarsdottir S, Cesaro S, Di Blasi R, Mikulska M, Rieger C, et al. Vaccination of haemopoietic stem cell transplant recipients: guidelines of the 2017 European Conference on Infections in Leukaemia (ECIL 7). Lancet Infect Dis. 2019 Feb 7;S1473-3099(18):30600–5.

30. Rubin LG, Levin MJ, Ljungman P, Davies EG, Avery R, Tomblyn M, et al. 2013 IDSA clinical practice guideline for vaccination of the immunocompromised host. Clinical Infectious Diseases. 2014;58(3):309–18.

31. Tomblyn M, Chiller T, Einsele H, Gress R, Sepkowitz K, Storek J, et al. Guidelines for preventing infectious complications among hematopoietic cell transplantation recipients: a global perspective. Biol Blood Marrow Transplant. 2009 Oct;15(10):1143–238.

32. Cordonnier C, Labopin M, Chesnel V, Ribaud P, De La Camara R, Martino R, et al. Randomized Study of Early versus Late Immunization with Pneumococcal Conjugate Vaccine after Allogeneic Stem Cell Transplantation. Clinical Infectious Diseases. 2009 May 15;48(10):1392–401.

33. Carpenter PA, Englund JA. How I vaccinate blood and marrow transplant recipients. Blood [Internet]. 2016 [cited 2019 Nov 20];127(23):blood-2015-12-550475. Available from: http://www.bloodjournal.org/content/early/2016/04/05/blood-2015-12-550475.abstract

34. Khawaja F, Papanicolaou G, Dadwal S, Pergam SA, Wingard JR, Boghdadly Z El, et al. Frequently Asked Questions on Coronavirus Disease 2019 Vaccination for Hematopoietic Cell Transplantation and Chimeric Antigen Receptor T-Cell Recipients From the American Society for Transplantation and Cellular Therapy and the American Society of Hematology. Transplant Cell Ther [Internet]. 2022 Oct [cited 2022 Nov 30]; Available from: https://pubmed.ncbi.nlm.nih.gov/36273782/

35. National Comprehensive Cancer Network. Recommendations of the NCCN COVID-19 Vaccination Advisory Committee [Internet]. 2021 [cited 2021 Jul 20]. Available from: https://www.nccn.org/docs/default-source/covid-19/2021_covid-19_vaccination_guidance_v2-0.pdf

36. Kampouri E, Hill JA, Dioverti V. COVID-19 after hematopoietic cell transplantation and chimeric antigen receptor (CAR)-T-cell therapy. Transpl Infect Dis [Internet]. 2023 [cited 2023 Dec 10];25 Suppl 1. Available from: https://pubmed.ncbi.nlm.nih.gov/37767643/

37. Reynolds G, Hall VG, Teh BW. Vaccine schedule recommendations and updates for patients with hematologic malignancy post-hematopoietic cell transplant or CAR T-cell therapy. Transpl Infect Dis [Internet]. 2023 [cited 2023 Dec 10];25 Suppl 1. Available from: https://pubmed.ncbi.nlm.nih.gov/37515788/

38. Mittal A, Solera JT, Ferreira VH, Kothari S, Kimura M, Pasic I, et al. Immunogenicity and Safety of Booster SARS-CoV-2 mRNA Vaccine Dose in Allogeneic Hematopoietic Stem Cell Transplantation Recipients. Transplant Cell Ther [Internet]. 2023 Nov 1 [cited 2023 Dec 10];29(11):706.e1–706.e7. Available from: https://pubmed.ncbi.nlm.nih.gov/37582470/

39. Nikoloudis A, Neumann IJ, Buxhofer-Ausch V, Machherndl-Spandl S, Binder M, Kaynak E, et al. Successful SARS-CoV-2 mRNA Vaccination Program in Allogeneic Hematopoietic Stem Cell Transplant Recipients-A Retrospective Single-Center Analysis. Vaccines (Basel) [Internet]. 2023 Oct 1 [cited 2023 Dec 10];11(10). Available from: https://pubmed.ncbi.nlm.nih.gov/37896938/

40. Hill JA, Martens MJ, Young JAH, Bhavsar K, Kou J, Chen M, et al. SARS-CoV-2 vaccination in the first year after allogeneic hematopoietic cell transplant: a prospective, multicentre, observational study. EClinicalMedicine [Internet]. 2023 May 1 [cited 2023 Dec 10];59. Available from: https://pubmed.ncbi.nlm.nih.gov/37128256/

41. Huang Y, Borisov O, Kee JJ, Carpp LN, Wrin T, Cai S, et al. Calibration of two validated SARS-CoV-2 pseudovirus neutralization assays for COVID-19 vaccine evaluation. Sci Rep. 2021;11(1):1–13.

42. Alter G, Yu J, Liu J, Chandrashekar A, Borducchi EN, Tostanoski LH, et al. Immunogenicity of Ad26.COV2.S vaccine against SARS-CoV-2 variants in humans. Nature. 2021 Jun 9;596(7871):268–72.

43. Carlson CS, Emerson RO, Sherwood AM, Desmarais C, Chung MW, Parsons JM, et al. Using synthetic templates to design an unbiased multiplex PCR assay. Nat Commun. 2013 Oct 25;4(2680).

44. Robins HS, Campregher P V., Srivastava SK, Wacher A, Turtle CJ, Kahsai O, et al. Comprehensive assessment of T-cell receptor β-chain diversity in αβ T cells. Blood. 2009 Nov 5;114(19):4099–107.

45. Zonozi R, Walters LC, Shulkin A, Naranbhai V, Nithagon P, Sauvage G, et al. T cell responses to SARS-CoV-2 infection and vaccination are elevated in B cell deficiency and reduce risk of severe COVID-19. Sci Transl Med [Internet]. 2023 Nov 29 [cited 2023 Dec 11];15(724):eadh4529. Available from: https://www.science.org/doi/10.1126/scitranslmed.adh4529

46. Bange EM, Han NA, Wileyto P, Kim JY, Gouma S, Robinson J, et al. CD8+ T cells contribute to survival in patients with COVID-19 and hematologic cancer. Nat Med [Internet]. 2021 Jul 1 [cited 2022 Feb 8];27(7):1280–9. Available from: https://www.mendeley.com/catalogue/8dbcb21d-a658-3d9a-8e71-6e031f3c5cc2/?ref=raven&dgcid=raven_md_suggest_email&dgcid=raven_md_suggest_mie_email

47. Khawaja F, Papanicolaou G, Dadwal S, Pergam SA, Wingard JR, Boghdadly Z El, et al. Frequently Asked Questions on Coronavirus Disease 2019 Vaccination for Hematopoietic Cell Transplant and Chimeric Antigen Receptor T-Cell Recipients From the American Society for Transplantation and Cellular Therapy and the American Society of Hematology. Transplant Cell Ther. 2022 Oct;

48. Ljungman P, Cordonnier C, Einsele H, Englund J, Machado CM, Storek J, et al. Vaccination of hematopoietic cell transplant recipients. Bone Marrow Transplant. 2009;44(8):521–6.

49. Hill JA, Seo S. How we prevent infections in patients receiving CD19-targeted chimeric antigen receptor T-cells for B-cell malignancies. Blood. 2020 Jun;

50. Sherman AC, Cheng CA, Swank Z, Zhou G, Li X, Issa NC, et al. Impact of Donor and Recipient SARS-CoV-2 Vaccination or Infection on Immunity after Hematopoietic Cell Transplantation. Transplant Cell Ther [Internet]. 2023 Feb [cited 2023 Mar 13]; Available from: https://pubmed.ncbi.nlm.nih.gov/36736784/

51. La Rosa C, Chiuppesi F, Park Y, Zhou Q, Yang D, Gendzekhadze K, et al. Functional SARS-CoV-2-specific T cells of donor origin in allogeneic stem cell transplant recipients of a T-cell-replete infusion: A prospective observational study. Front Immunol [Internet]. 2023 [cited 2024 Jan 7];14. Available from: https://pubmed.ncbi.nlm.nih.gov/36936918/

