## Supplement for "SARS-CoV-2 vaccination in the first year after hematopoietic cell transplant or chimeric antigen receptor T cell therapy: A prospective, multicenter, observational study (BMT CTN 2101)"

[**Table S1.** *A priori* power to detect a 25% difference in immunogenicity rates between 6-12 and <6 month cohorts with 41 and 112 evaluable patients within each cohort.	14](#_Toc156984964)

[**Table S2.** *Post hoc* power to detect a 25% difference in immunogenicity rates between 4-12 and <4 month subgroups per cohort.	15](#_Toc156984965)

[**Table S3.** Comparisons of anti-S IgG and neutralizing antibodies between the <4 month and 4-12 month vaccination timing subgroup at each time point within each cellular therapy cohort, based on the data depicted in Figure 1A and 1B.	16](#_Toc156984966)

[**Table S4.** Comparisons of anti-S IgG and neutralizing antibodies between subsequent time points within each vaccination timing subgroup (<4 month and 4-12 month) and cellular therapy cohort, based on the data depicted in Figure 1A and 1B.	17](#_Toc156984967)

### LIST OF INVESTIGATORS

Paul Armistead, University of North Carolina Medical Center, Chapel Hill, NC; Jo-Anne Young, University of Minnesota, Minneapolis, MN; Jan Cerny, UMass Memorial Medical Center, Worcester, MA; Edward Copelan, Levine Cancer Institute, Charlotte, NC; David Epstein, Stanford Hospital, Stanford, CA; Uroosa Ibrahim, Mount Sinai Hospital, New York, NY; Mehdi Hamadani, Medical College of Wisconsin, Milwaukee, WI; Nancy Hardy, University of Maryland, College Park, MD; Joshua Hill, Fred Hutchinson Cancer Research Center, Seattle, WA; Kent Holland, Northside Hospital, Atlanta, GA; Mitch Horwitz, Duke University Hospital, Durham, NC; Dianna Howard, Wake Forest Baptist, Winston-Salem, NC; Robert Krance, Baylor College of Medicine, Houston, TX; Richard Maziarz, Oregon Health & Science University, Portland, OR; Philip McCarthy, Roswell Park Comprehensive Cancer Center, Buffalo, NY; John McCarty, Virginia Commonwealth University, Richmond, VA; Joseph McGuirk, University of Kansas, Lawrence, KS; Miguel Perales, Memorial Sloan Kettering Cancer Center, New York, NY; Ron Sobecks, Cleveland Clinic, Cleveland, OH; Lauren Veltri, West Virginia University, Morgantown, WV; Ned Waller, Emory University Hospital, Atlanta, GA; Peter Westervelt, Barnes-Jewish Hospital, Washington University, St. Louis, MO.

### METHODS

**Data Source and Collection**

The Center for International Blood and Marrow Transplant Research (CIBMTR) is a research consortium consisting of over 500 transplant centers internationally. Through a collaboration between the Medical College of Wisconsin and the National Marrow Donor Program, patient and outcomes data from these centers are collected and analyzed. Central auditing of the data is performed to ensure consistency and quality. The CIBMTR collects comprehensive demographic and clinical data prior to transplantation, at 100 days (D100), 6 months (D180), and 1 year after transplantation and annually thereafter. All patients included in this study gave written consent to participate in the CIBMTR Research Database and to have their data included in observational research. Participants also signed a study-specific consent for additional data and blood sample collection specific to SARS-CoV-2 vaccination and follow up as detailed in the manuscript. This study was approved by the institutional review boards of the Medical College of Wisconsin and the National Marrow Donor Program.

Data collected at baseline and each time point included demographics and cellular therapy characteristics, medications (including for prevention or treatment of COVID-19), vaccine type, possible vaccine-related grade ≥3 adverse events according to NCI CTCAE Version 5.0, prior or new graft-versus-host disease (GVHD) and severity, and incident SARS-CoV2 infection.

The study was conducted under two CIBMTR repository protocols (“Protocol for a Research Database for Hematopoietic Cell Transplantation, Other Cellular Therapies and Marrow Toxic Injuries” and “Protocol for a Research Sample Repository for Hematopoietic Cell Transplantation, Other Cellular Therapies and Marrow Toxic Injuries”) that centers had ongoing institutional review board (IRB) approval for. The study-specific informed consent and assent for each center was approved by the National Marrow Donor Program (NMDP) single IRB.

**Participants and Study Design**

Prior SARS-CoV-2 infection, pre-hematopoietic SARS-CoV-2 vaccination, receipt of immunoglobulin replacement therapy, and receipt of prophylactic tixagevimab-cilgavimab were not exclusionary.

**Testing**

***Binding and neutralizing antibodies***. Total SARS-CoV-2 anti-spike receptor binding domain IgG was tested for in serum using the Roche Elecsys Anti-SARS-CoV-2 S electrochemiluminescence immunoassay (ECLIA).

Neutralizing antibody activity was measured in a FDA approved assay as previously described.^1^ Briefly, the assay uses lentiviral particles pseudotyped with full-length SARS-CoV-2 Spike protein and containing a firefly luciferase (Luc) reporter gene for quantitative measurements of infection by relative luminescence units (RLU). The backbone vector used in pseudovirus creation, F-lucP.CNDO∆U3, encodes the HIV genome with firefly luciferase replacing the HIV *env* gene. A codon-optimized version of the full-length spike gene of the Wuhan-1 SARS-CoV-2 strain (MN908947.3) (GenScript) was cloned into the Monogram proprietary *env* expression vector, pCXAS-PXMX. The D614G spike mutation was introduced into the original Wuhan sequence by site-directed mutagenesis. Pseudovirus infectivity was screened at multiple dilutions using HEK293 cells transiently transfected with ACE2 and TMPRSS2 expression vectors. RLUs were adjusted to ~ 50,000 for use in the neutralization assay. Neutralization was performed in 96-well plates by incubating pseudovirus with 10 serial three-fold dilutions of serum samples for one hour at 37 °C. Serum samples were heat-inactivated for 60 min at 56 °C prior to assay. The dilution series was based on a 1:20 starting dilution which was reported as 1:40 after addition of virus. Neutralization titers represent the inhibitory dilution (ID) of serum samples at which RLUs were reduced by 50% (ID50) compared to virus control wells (no serum wells). The units are 1/dilution, so an ID50 of 134 is a 1:134 dilution of the test serum at the 50% inhibition point.

Time points at which anti-S IgG was not detected (<0.4 U/mL) were not tested for neutralizing antibodies. Anti-S IgG values >0.8 units per milliliter (U/mL) and neutralizing titers ≥40 inhibitory dose (ID50) (reciprocal of the sample dilution required to reduce relative luminescence units by 50%) were considered positive as previously described.[32] Titers below the limit of detection (LOD) were assigned a value of one-half the LOD.

***Multiparametric Flow Cytometric Analysis*.** Cryopreserved peripheral blood mononuclear cells (PBMCs) were thawed, washed and resuspended in PBS, then incubated with Human TruStain FcX Fc receptor blocking solution (Biolegend) and Live/DEAD Fixable Blue Dead Cell Stain (Invitrogen) according to the manufacturers’ specifications for 20 minutes at room temperature (RT), protected from light. The cells were washed once in RPMI 1640 no phenol red + 4% FBS +0.01% sodium-azide and incubated with the antibody mix for 20 minutes at RT in the dark in the presence of Brilliant Staining Buffer (BD). The cells were washed, resuspended in 0.5% paraformaldehyde/PBS, and immediately acquired using a Cytek Aurora 5L flow cytometer (Cytek). The optimal concentration of all antibodies used in the study was defined by titration. Further information about the antibodies can be found in **Table S11**. For analysis, single-cell data was clustered using the FlowSOM R package and labeled using the Ek'Balam algorithm.^2,3^ Cell subset definitions were used as previously described.^4,5^ Cluster labeling, method implementation, and visualization were done through the Astrolabe Cytometry Platform (Astrolabe Diagnostics, Inc.).

**Statistical Analysis**

Antibody results from samples collected within six months of receipt of SARS-CoV-2-specific monoclonal antibodies were excluded and separately described.

We computed an *a priori* power calculation of the proportion of participants with immunogenicity between patients vaccinated 6-12 months versus <6 months after allogeneic HCT using a two sample Z test of the difference in proportions at a significance level of 5%. Immunogenicity for the a priori power calculation at the time of protocol development was defined as a ≥4-fold rise in anti-S IgG. The study was designed to provide at least 81% power to detect a 25% difference in immunogenicity response rates between timing cohorts by enrolling at least 118 and 43 patients to the <6 month and ≥6-12 month cohorts, respectively, assuming a 5% dropout rate (**Table S1**). Unequal allocation to the cohorts was expected based on the numbers of patients anticipated to enroll in these cohorts given clinical practice patterns at the time.

We also computed a *post-hoc* power calculation based on the sample sizes actually observed in the <4-month and 4-12 month timing cohorts (**Table S2**). The observed sample sizes for the allogeneic HCT, autologous HCT, and CAR-T therapy cohorts provided respective power levels of at least 93%, 81%, and 45% to detect a 25% difference in immunogenicity rates between the <4 month and 4-12 month timing groups.

Anti-S IgG, neutralizing antibodies, and TCR results are displayed in box-and-whisker plots and compared using nonparametric Wilcoxon rank sum tests that are robust to features such as skewness and high dispersion which may arise in immunogenicity endpoints.

To adjust for imbalances in baseline variables between timing cohorts, propensity scores for the likelihood of being in the <4-month cohort were constructed using logistic regression with stepwise variable selection. Variables from Tables 1 and 2 with p-values <0.05 were included in the model. A propensity-adjusted analysis compared positive anti-S IgG responses at the post-V2, post-V3, and end-of-study time points between the <4 month and 4–12-month cohorts. For each timing cohort, inverse probability weights were constructed from the reciprocal of the propensity of being included in the cohort; weighted response proportions and their standard errors were computed and used to obtain point estimates and 99% CIs for the propensity-adjusted difference in response rates.

Logistic and linear regression models evaluated the impact of vaccination timing on anti-S IgG positive responses with adjustment for other covariates determined by bidirectional stepwise selection, with vaccine timing forced into the model and p-values <0.05 as the criterion for inclusion of covariates. All participants were included in analyses as relevant (e.g., participants who did not receive a second vaccine were not excluded). Analyses were performed using SAS Version 9.4 and R version 4.2. A p-value threshold of 0.01 was used to determine significance for all statistical comparisons to account for multiple comparisons.

### SUPPLEMENTARY FIGURES

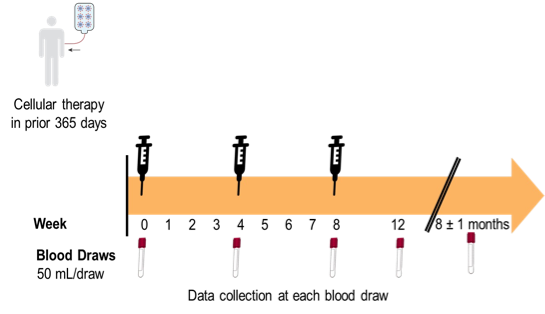

#### **Figure S1**. Enrollment and blood collection schema.

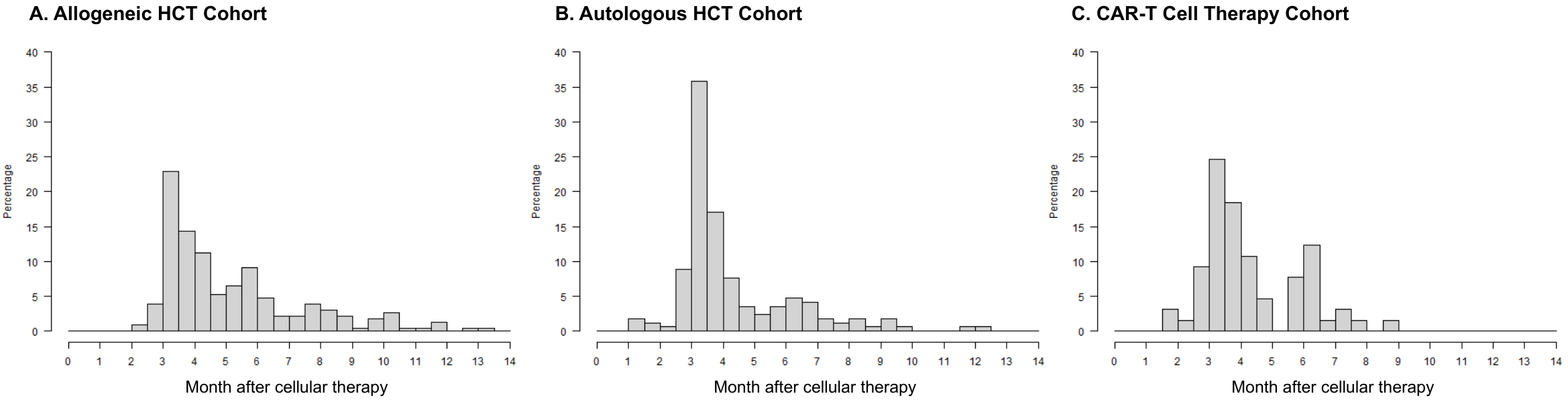

**Figure S2.** Histogram of the month of first SARS-CoV-2 vaccination after cellular therapy per therapy type. The median (interquartile range [IQR]) time to vaccination was 4.3 months (3.5-6.1) after allogeneic HCT, 3.6 months (3.3-4.9) after autologous HCT, and 3.9 months (3.3-5.8) after CAR-T cell therapy. Three participants were enrolled within 12 months but received their first vaccine >12 months after treatment, 2 in the allogeneic HCT cohort (12.8, 13.4 months) and 1 in the autologous HCT cohort (12.1 months).

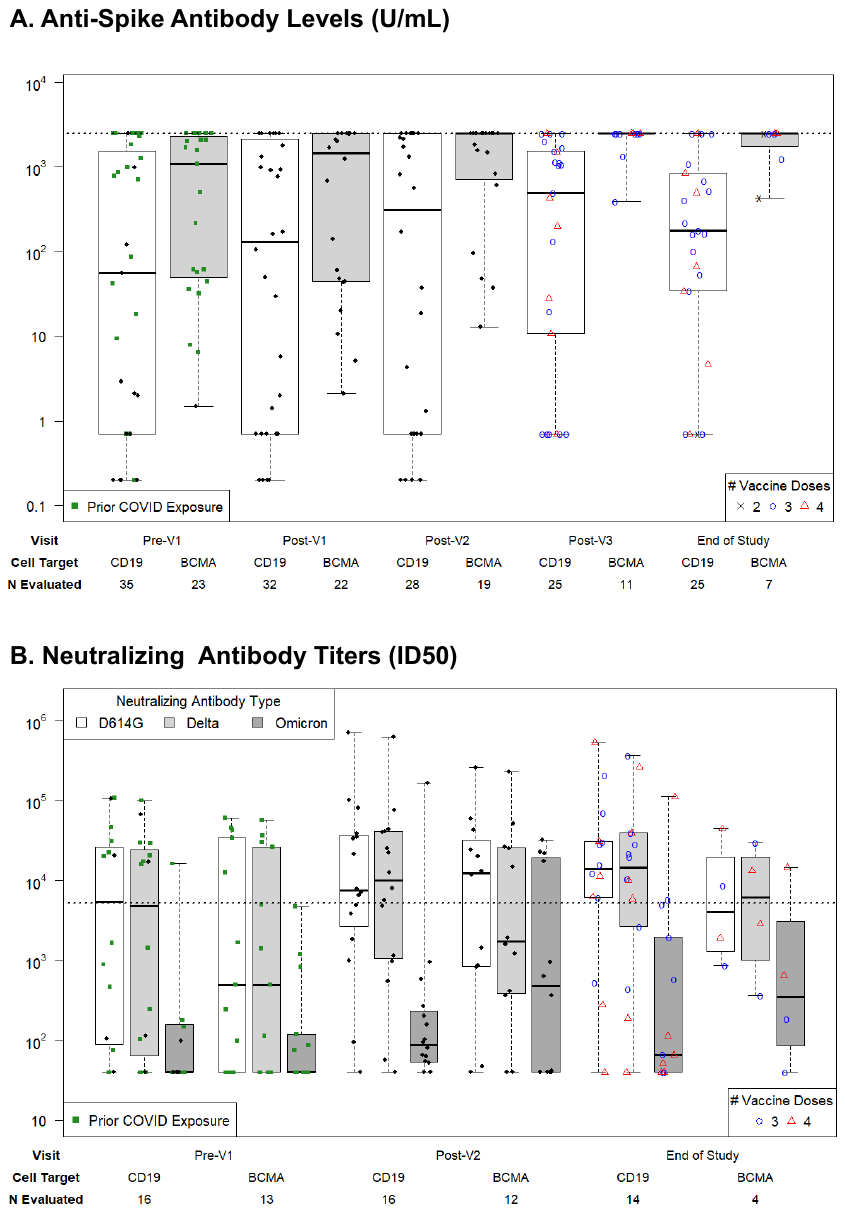

#### **Figure S3.** Longitudinal SARS-CoV-2 anti-Spike IgG titers and neutralizing antibody titers among CAR-T cell therapy recipients stratified by CAR-T cell target.

**A)** SARS-CoV-2 anti-S IgG titers per time point. The horizontal dotted line indicates the threshold for a positive response, defined as anti-S IgG >2,500 U/mL as determined from a ROC curve analysis; this was also the upper limit of quantitation for the assay. **B)** SARS-CoV-2 neutralizing antibody titers (ID50); ID50 is defined as the reciprocal of the sample dilution required to reduce relative luminescence units by 50%. The horizontal dotted line shows the median neutralizing antibody level (5,274 ID50) achieved in a healthy cohort vaccinated with two doses of mRNA-1273 (Moderna) in a clinical trial and tested with the same assay and defined here as a positive response. In panels A and B, ‘prior COVID exposure’ (green squares) indicates data in the first two time points from participants with a known prior SARS-CoV-2 infection, prior SARS-Cov-2 vaccination in the participant or hematopoietic cell donor, or positive anti-N IgG assay at baseline. Results are depicted on a log_10_ scale. Time points tested within 6 months of receipt of tixagevimab-cilgavimab (Evusheld) were excluded.

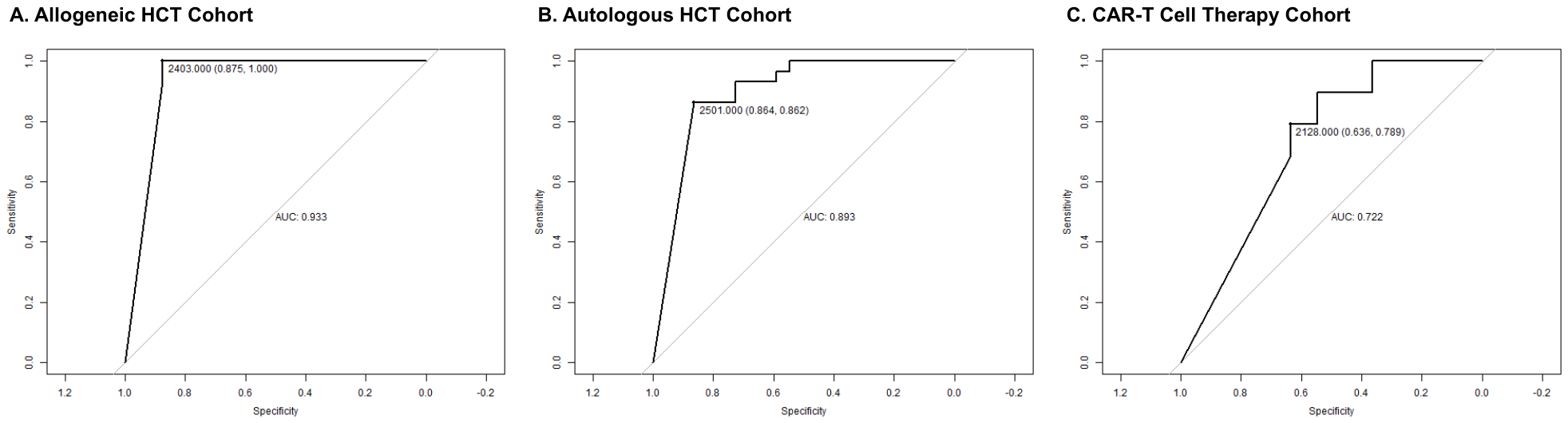

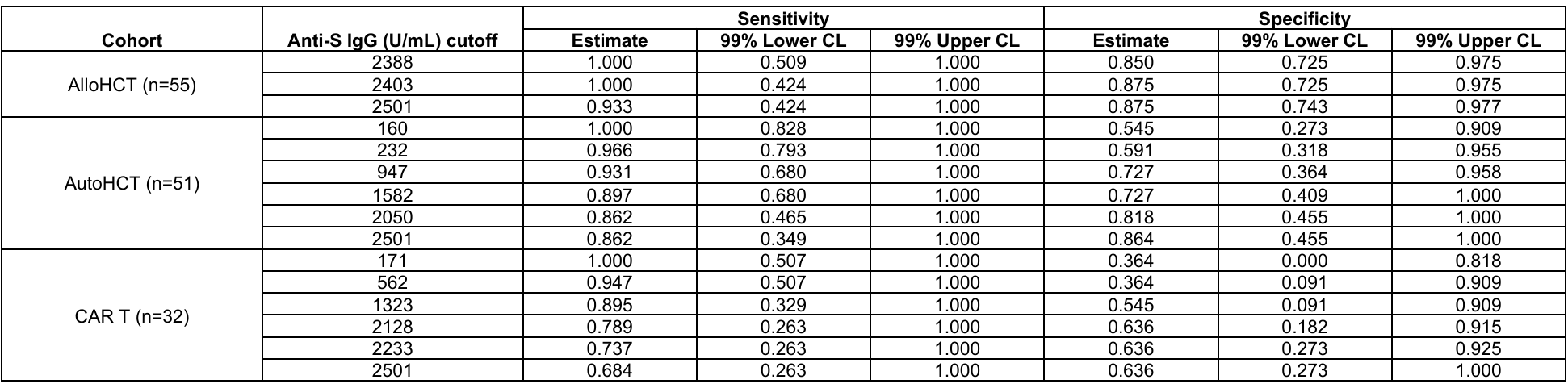

#### **Figure S4.** Receiver operating characteristic (ROC) curves of SARS-CoV-2 anti-S IgG titers as a predictor for neutralizing antibodies.

Receiver operating characteristic (ROC) curves of SARS-CoV-2 anti-S IgG titers as a predictor for neutralizing antibodies are demonstrated for the post-V2 time point, based on the median level of neutralizing antibodies (5,274 ID50) achieved in a healthy cohort vaccinated with two doses of mRNA-1273 (Moderna) in a clinical trial and tested with the same assay.^30^ The allogeneic HCT, autologous HCT, and CAR-T cell therapy cohorts are depicted in Panels A, B, and C, respectively. Patients who received tixagevimab-cilgavimab (Evusheld) within the prior six months were excluded for all analyses. The data are depicted in a Table below the panels.

CL indicates confidence limit; AUC, area under the curve.

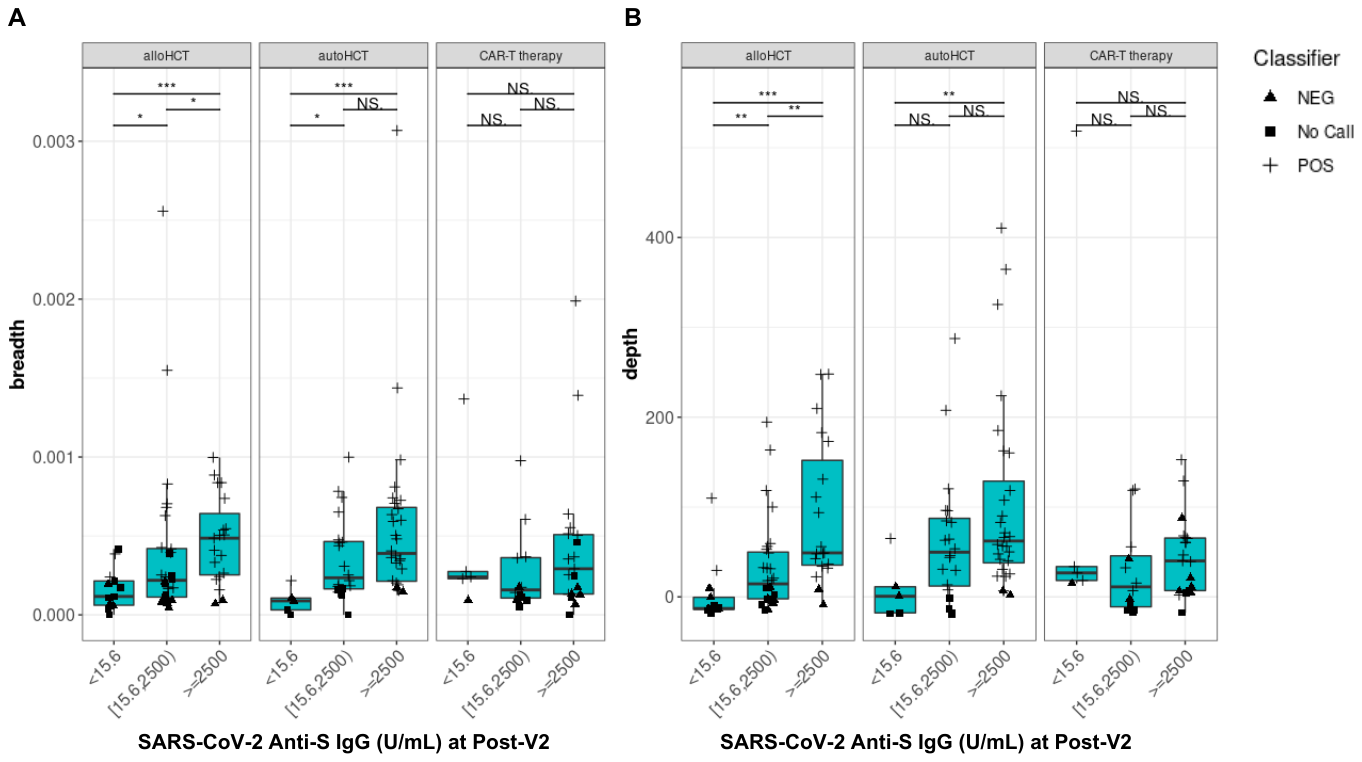

#### **Figure S5.** SARS-CoV-2-specific T cell receptor (TCR) variable beta chain sequencing results in a subgroup of 151 participants.

Quantitative values of SARS-CoV-2 TCR breadth (A) and depth (B) at the post-V2 time point in categories of negative, any detectable, or positive SARS-CoV-2 anti-S IgG titers. Any detectable antibody (≥15.6) was based on a threshold determined to be predictive of detection of neutralizing antibodies at any level. Individuals with a positive T-Detect at the pre-V1 time point were excluded.

NS indicates not significant; *, ≤0.05; **, ≤0.01; ***, ≤0.001.

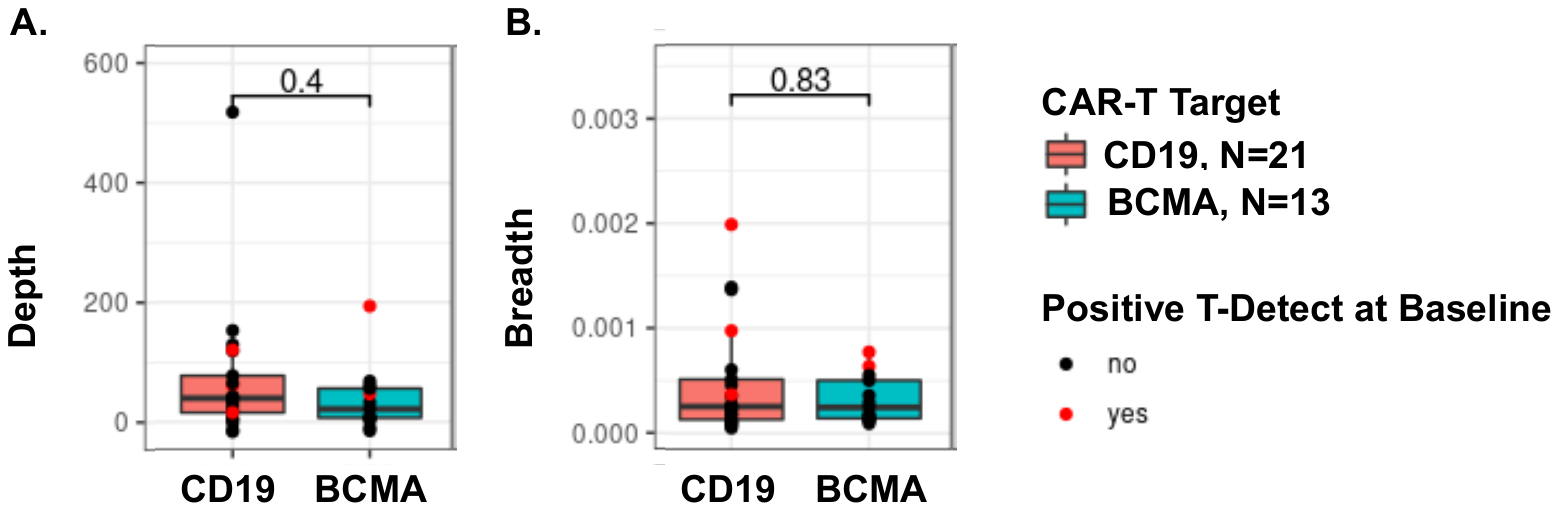

#### **Figure S6.** SARS-CoV-2-specific T cell receptor (TCR) variable beta chain sequencing results stratified by CAR-T cell target at the post-V2 time point.

Quantitative values of SARS-CoV-2 TCR depth (A) and breadth (B) at the post-V2 time point in categories of CD19 versus BCMA CAR-T cell target. The results were similar when excluding participants with a postivie T-Detect at baseline. Qualitative results were also similar, with a positive T-Detect in 60% and 61.5% of CD19 and BCMA-CAR-T cell recipients, respectively.

**SUPPLEMENTARY TABLES**

| **Table S1.** *A priori* power to detect a 25% difference in immunogenicity rates between 6-12 and <6 month cohorts with 41 and 112 evaluable patients within each cohort. | |
| --- | --- |
| **Difference in Response Rates (6-12 mos. vs. < 6 mos.)** | **Power** |
| 90% vs. 65% or 10% vs. 35% | 92.8% |
| 80% vs. 55% or 20% vs. 45% | 85.4% |
| 70% vs. 45% or 30% vs. 55% | 81.7% |
| 60% vs. 35% or 40% vs. 65% | 81.6% |

| **Table S2.** *Post hoc* power to detect a 25% difference in immunogenicity rates between 4-12 and <4 month subgroups per cohort. | |
| --- | --- |
| **Difference in Response Rates (4-12 months vs. < 4 months)** | **Power** |
| Allogeneic HCT (N = 119 versus N = 76) | |
| 90% vs. 65% or 10% vs. 35% | 98.7% |
| 80% vs. 55% or 20% vs. 45% | 96.0% |
| 70% vs. 45% or 30% vs. 55% | 93.8% |
| 60% vs. 35% or 40% vs. 65% | 93.3% |
| Autologous HCT (N = 50 versus N = 87) | |
| 90% vs. 65% or 10% vs. 35% | 92.9% |
| 80% vs. 55% or 20% vs. 45% | 85.8% |
| 70% vs. 45% or 30% vs. 55% | 82.0% |
| 60% vs. 35% or 40% vs. 65% | 81.5% |
| CAR-T cell therapy cohort (N = 26 versus N = 28) | |
| 90% vs. 65% or 10% vs. 35% | 59.4% |
| 80% vs. 55% or 20% vs. 45% | 49.7% |
| 70% vs. 45% or 30% vs. 55% | 45.6% |
| 60% vs. 35% or 40% vs. 65% | 45.0% |

| **Table S3.** Comparisons of anti-S IgG and neutralizing antibodies between the <4 month and 4-12 month vaccination timing subgroup at each time point within each cellular therapy cohort, based on the data depicted in Figure 1A and 1B. | | | | | |
| --- | --- | --- | --- | --- | --- |
| **Test type** | **Time point** | | | | |
|  | **Pre-V1** | **Post-V1** | **Post-V2** | **Post-V3** | **End-of-study** |
| Allogeneic HCT | | | | | |
| Anti-S IgG | 0.002 | 0.005 | 0.191 | 0.778 | 0.843 |
| Neutralizing, Wuhan D614G | 1.000 | NA | 0.784 | NA | 0.571 |
| Neutralizing, Delta B.1.617.2 | 0.807 | NA | 0.798 | NA | 0.797 |
| Neutralizing, Omicron B.1.1.529 | 0.268 | NA | 0.274 | NA | 0.718 |
| Autolgous HCT | | | | | |
| Anti-S IgG | 0.184 | 0.648 | 0.444 | 0.730 | 0.344 |
| Neutralizing, Wuhan D614G | 0.017 | NA | 0.052 | NA | 0.447 |
| Neutralizing, Delta B.1.617.2 | 0.032 | NA | 0.079 | NA | 0.420 |
| Neutralizing, Omicron B.1.1.529 | 0.021 | NA | 0.126 | NA | 0.661 |
| CAR-T cell therapy | | | | | |
| Anti-S IgG | 0.117 | 0.484 | 0.279 | 0.282 | 0.156 |
| Neutralizing, Wuhan D614G | 0.100 | NA | 0.704 | NA | 0.186 |
| Neutralizing, Delta B.1.617.2 | 0.141 | NA | 0.403 | NA | 0.345 |
| Neutralizing, Omicron B.1.1.529 | 0.544 | NA | 0.674 | NA | 0.772 |
| Wilcoxon rank sum test p-values are shown comparing median antibody levels between cohorts. Participants who received tixagevimab-cilgavimab (Evusheld) within 6 months prior to a visit were excluded. | | | | | |

| **Table S4.** Comparisons of anti-S IgG and neutralizing antibodies between subsequent time points within each vaccination timing subgroup (<4 month and 4-12 month) and cellular therapy cohort, based on the data depicted in Figure 1A and 1B. | | | | |
| --- | --- | --- | --- | --- |
| **Timing Cohort and test type** | **Time point** | | | |
|  | **Pre-V1 vs. Post-V1** | **Post-V1 vs. Post-V2^a^** | **Post-V2 vs. Post-V3** | **Post-V3 vs. end-of-study^b^** |
| **Allogeneic HCT** | | | | |
| **<4 months** |  |  |  |  |
| Anti-S IgG | 0.040 | <0.001 | <0.001 | 0.047 |
| Neutralizing, Wuhan D614G | NA | 0.006 | NA | 0.131 |
| Neutralizing, Delta B.1.617.2 | NA | 0.005 | NA | 0.131 |
| Neutralizing, Omicron B.1.1.529 | NA | 0.021 | NA | 0.080 |
| **4-12 months** |  |  |  |  |
| Anti-S IgG | 0.163 | <0.001 | <0.001 | 0.985 |
| Neutralizing, Wuhan D614G | NA | 0.001 | NA | <0.001 |
| Neutralizing, Delta B.1.617.2 | NA | 0.002 | NA | <0.001 |
| Neutralizing, Omicron B.1.1.529 | NA | 0.722 | NA | <0.001 |
| **Autologous HCT** | | | | |
| **<4 months** |  |  |  |  |
| Anti-S IgG | 0.045 | 0.011 | 0.002 | 0.103 |
| Neutralizing, Wuhan D614G | NA | <0.001 | NA | 0.834 |
| Neutralizing, Delta B.1.617.2 | NA | <0.001 | NA | 0.780 |
| Neutralizing, Omicron B.1.1.529 | NA | 0.004 | NA | 0.235 |
| **4-12 months** |  |  |  |  |
| Anti-S IgG | <0.001 | 0.071 | 0.013 | 0.563 |
| Neutralizing, Wuhan D614G | NA | 0.030 | NA | 0.623 |
| Neutralizing, Delta B.1.617.2 | NA | 0.046 | NA | 0.515 |
| Neutralizing, Omicron B.1.1.529 | NA | <0.001 | NA | 0.065 |
| **CAR-T cell therapy** | | | | |
| **<4 months** |  |  |  |  |
| Anti-S IgG | 0.920 | 0.268 | 0.141 | 0.097 |
| Neutralizing, Wuhan D614G | NA | 1.000 | NA | 0.129 |
| Neutralizing, Delta B.1.617.2 | NA | 0.685 | NA | 0.426 |
| Neutralizing, Omicron B.1.1.529 | NA | 0.359 | NA | 0.447 |
| **4-12 months** |  |  |  |  |
| Anti-S IgG | 0.808 | 0.237 | 0.706 | 0.351 |
| Neutralizing, Wuhan D614G | NA | 0.205 | NA | 0.262 |
| Neutralizing, Delta B.1.617.2 | NA | 0.289 | NA | 0.262 |
| Neutralizing, Omicron B.1.1.529 | NA | 0.021 | NA | 0.554 |
| Wilcoxon signed rank test p-values are shown, testing whether median changes in antibody levels between visits differ from 0. Participants who received tixagevimab-cilgavimab (Evusheld) within 6 months prior to a visit were excluded.  ^a^For neutralizing antibodies, the comparison is between pre-V1 and post-V2.  ^b^For neutralizing antibodies, the comparison is between post-V2 and end-of-study. | | | | |

| **Table S5.** Qualitative T-cell responses based on a positive T-Detect assay. | | | |
| --- | --- | --- | --- |
|  | **Pre-V1** | **Post-V2** | **End-Of-Study**^a^ |
| **Allogeneic HCT Cohort** | | | |
| **Positive** | 3 (5.0%) | 33 (55.0%) | 32 (57.1%) |
| **Negative** | 33 (55.0%) | 14 (23.3%) | 18 (32.1%) |
| **No call** | 24 (40.0%) | 13 (21.7%) | 6 (10.7%) |
| **Autologous HCT Cohort** | | | |
| **Positive** | 12 (22.6%) | 43 (81.1%) | 40 (83.3%) |
| **Negative** | 31 (58.5%) | 4 (7.5%) | 2 (4.2%) |
| **No call** | 10 (18.9%) | 6 (11.3%) | 6 (12.5%) |
| **CAR-T Cell Therapy Cohort** | | | |
| **Positive** | 11 (28.9%) | 23 (60.5%) | 18 (58.1%) |
| **Negative** | 15 (39.5%) | 8 (21.1%) | 6 (19.4%) |
| **No call** | 12 (31.6%) | 7 (18.4%) | 7 (22.6%) |
| ^a^Inclusive of participants with a positive result at the Pre-V1 time point. | | | |

| **Table S6.** Adjusted logistic regression models of the association of baseline and time-dependent variables with an observed positive SARS-CoV-2 anti-S IgG titer. | | | | | |
| --- | --- | --- | --- | --- | --- |
| **Variable^a^** | **Category** | **N** | **Odds Ratio** | **99% CI** | **p-value** |
| **Post-V2 time point** | | | | | |
| Cellular therapy and vaccine timing categories^b^ | AlloHCT, < 4 Months | 76 | 1.000 | - | 0.0950 (5df) |
|  | AlloHCT, 4 - 12 Months | 119 | 0.507 | (0.211, 1.218) | 0.0459 |
|  | AutoHCT, < 4 Months | 87 | 0.546 | (0.195, 1.528) | 0.1297 |
|  | AutoHCT, 4 - 12 Months | 50 | 1.060 | (0.353, 3.182) | 0.8915 |
|  | CAR-T, < 4 Months | 28 | 1.073 | (0.267, 4.314) | 0.8958 |
|  | CAR-T, 4 - 12 Months | 26 | 0.307 | (0.069, 1.372) | 0.0421 |
| Recipient SARS-CoV-2 infection pre-enrollment | No | 328 | 1.000 | - |  |
|  | Yes | 58 | 4.225 | (1.755, 10.169) | < 0.0001 |
| Recipient SARS-CoV-2 vaccination pre-enrollment | No | 233 | 1.000 | - |  |
|  | Yes | 153 | 3.025 | (1.502, 6.096) | < 0.0001 |
| B cell and T cell counts measured pre-enrollment | No | 109 | 1.000 | - |  |
|  | Yes | 277 | 0.598 | (0.293, 1.220) | 0.0634 |
| Log_10_ B cell count in measured patients^c^ | - | 386 | 2.093 | (1.227, 3.570) | 0.0004 |
| Log_10_ T cell count in measured patients^c^ | - | 386 | 1.981 | (0.976, 4.021) | 0.0129 |
| **Post-V3 time point** | | | | | |
| Vaccine timing cohort^b^ | AlloHCT, < 4 Months | 54 | 1.000 | - | 0.0257 (5df) |
|  | AlloHCT, 4 - 12 Months | 69 | 1.129 | (0.414, 3.080) | 0.7560 |
|  | AutoHCT, < 4 Months | 60 | 1.923 | (0.642, 5.759) | 0.1247 |
|  | AutoHCT, 4 - 12 Months | 31 | 1.632 | (0.431, 6.184) | 0.3434 |
|  | CAR-T, < 4 Months | 21 | 0.768 | (0.182, 3.241) | 0.6365 |
|  | CAR-T, 4 - 12 Months | 21 | 0.255 | (0.056, 1.165) | 0.0205 |
| B cell and T cell counts measured pre-enrollment | No | 71 | 1.000 | - |  |
|  | Yes | 185 | 0.570 | (0.246, 1.323) | 0.0855 |
| Log_10_ B cell count in measured patients^c^ | - | 256 | 1.884 | (1.035, 3.429) | 0.0064 |
| **End-of-study time point** | | | | | |
| Cellular therapy and vaccine timing categories^b^ | AlloHCT, < 4 Months | 46 | 1.000 | - | 0.0081 (5df) |
|  | AlloHCT, 4 - 12 Months | 83 | 1.086 | (0.377, 3.124) | 0.8414 |
|  | AutoHCT, < 4 Months | 59 | 1.004 | (0.326, 3.090) | 0.9930 |
|  | AutoHCT, 4 - 12 Months | 44 | 1.446 | (0.400, 5.227) | 0.4598 |
|  | CAR-T, < 4 Months | 18 | 0.388 | (0.074, 2.034) | 0.1412 |
|  | CAR-T, 4 - 12 Months | 18 | 0.102 | (0.016, 0.658) | 0.0016 |
| Recipient SARS-CoV-2 infection pre-enrollment | No | 227 | 1.000 | - |  |
|  | Yes | 41 | 3.271 | (0.999, 10.706) | 0.0101 |
| HCT-CI | 0 | 60 | 1.000 | - |  |
|  | ≥1 | 208 | 0.355 | (0.135, 0.935) | 0.0059 |
| B cell and T cell counts measured pre-enrollment | No | 73 | 1.000 | - |  |
|  | Yes | 195 | 0.537 | (0.224, 1.289) | 0.0673 |
| Log_10_ B cell count in measured patients^c^ | - | 268 | 1.766 | (0.976, 3.193) | 0.0135 |
| N indicates number; CI indicates confidence interval.  Response is defined as an anti-Spike IgG level >2,500 U/mL. A stepwise variable selection procedure was used to determine covariates to include in the model, with a p-value < 0.05 as the criterion for inclusion. Cellular therapy and vaccine initation timing were included in all models. The displayed variables were the only variables retained in the adjusted models. Potential interactions were evaluated between covariates. Patients who received Evusheld before the visit’s sample collection were excluded from the analysis. ^a^Covariates considered in models included baseline and time dependent variables from Tables 1 and 2. Time-dependent variables were considered in models at baseline as well as at the time point preceding each endpoint.  ^b^Adjusted response rate estimates and confidence intervals were constructed for each cellular therapy and vaccine timing subgroup and compared between timing groups within a cell therapy cohort (e.g., among allogeneic HCT only, autologous HCT only, and CAR-T cell therapy only). No significant differences were observed.  ^c^Median B cell count was 84.19 cells/uL; median T cell count was 272.35 cells/uL. | | | | | |

| **Table S7.** Adjusted linear regression models of the association of baseline and time-dependent variables with SARS-CoV-2 anti-S IgG titer (analyzed as a continuous variable per log_10_ increase). | | | | | |
| --- | --- | --- | --- | --- | --- |
| **Variable^a^** | **Category** | **N** | **Estimate** | **99% CI** | **p-value** |
| **Post-V2 time point** | | | | | |
| Cellular therapy and vaccine timing categories^b^ | AlloHCT, < 4 Months | 76 | 0.000 | - | 0.4658 (5df) |
|  | AlloHCT, 4 - 12 Months | 119 | -0.170 | (-0.552, 0.213) | 0.2535 |
|  | AutoHCT, < 4 Months | 87 | -0.135 | (-0.559, 0.290) | 0.4131 |
|  | AutoHCT, 4 - 12 Months | 50 | 0.034 | (-0.443, 0.511) | 0.8551 |
|  | CAR-T, < 4 Months | 28 | -0.274 | (-0.898, 0.349) | 0.2570 |
|  | CAR-T, 4 - 12 Months | 26 | -0.378 | (-0.994, 0.238) | 0.1142 |
| Recipient SARS-CoV-2 infection pre-enrollment | No | 328 | 0.000 | - |  |
|  | Yes | 58 | 0.527 | (0.158, 0.897) | 0.0002 |
| Recipient SARS-CoV-2 vaccination pre-enrollment | No | 233 | 0.000 | - |  |
|  | Yes | 153 | 0.666 | (0.354, 0.978) | < 0.0001 |
| B cell and T cell counts measured pre-enrollment | No | 109 | 0.000 | - |  |
|  | Yes | 277 | -0.163 | (-0.465, 0.139) | 0.1648 |
| Log_10_ B cell count in measured patients^c^ | - | 386 | 0.482 | (0.279, 0.685) | < 0.0001 |
| **Post-V3 time point** | | | | | |
| Vaccine timing cohort^b^ | AlloHCT, < 4 Months | 54 | 0.000 | - | 0.0147 (5df) |
|  | AlloHCT, 4 - 12 Months | 69 | 0.064 | (-0.366, 0.494) | 0.7016 |
|  | AutoHCT, < 4 Months | 60 | 0.135 | (-0.332, 0.602) | 0.4569 |
|  | AutoHCT, 4 - 12 Months | 31 | 0.071 | (-0.473, 0.615) | 0.7352 |
|  | CAR-T, < 4 Months | 21 | -0.402 | (-1.056, 0.252) | 0.1130 |
|  | CAR-T, 4 - 12 Months | 21 | -0.645 | (-1.282, -0.009) | 0.0090 |
| Recipient SARS-CoV-2 infection pre-enrollment | No | 220 | 0.000 | - |  |
|  | Yes | 36 | 0.527 | (0.158, 0.897) | 0.0002 |
| Recipient SARS-CoV-2 vaccination pre-enrollment | No | 145 | 0.000 | - |  |
|  |  | 111 | 0.651 | (0.285, 1.070) | < 0.0001 |
| B cell and T cell counts measured pre-enrollment | No | 71 | 0.000 | - |  |
|  | Yes | 185 | -0.163 | (-0.465, 0.139) | 0.1648 |
| Log_10_ B cell count in measured patients^c^ | - | 256 | 0.482 | (0.279, 0.685) | < 0.0001 |
| **End-of-study time point** | | | | | |
| Cellular therapy and vaccine timing categories^b^ | AlloHCT, < 4 Months | 46 | 0.000 | - | < 0.0001 (5df) |
|  | AlloHCT, 4 - 12 Months | 83 | -0.148 | (-0.621, 0.326) | 0.4223 |
|  | AutoHCT, < 4 Months | 59 | -0.262 | (-0.835, 0.312) | 0.2398 |
|  | AutoHCT, 4 - 12 Months | 44 | -0.197 | (-0.762, 0.368) | 0.3684 |
|  | CAR-T, < 4 Months | 18 | -0.905 | (-1.677, -0.133) | 0.0025 |
|  | CAR-T, 4 - 12 Months | 18 | -1.384 | (-2.138, -0.631) | < 0.0001 |
| Recipient SARS-CoV-2 infection pre-enrollment | No | 227 | 0.000 | - |  |
|  | Yes | 41 | 0.460 | (0.028, 0.892) | 0.0060 |
| Recipient SARS-CoV-2 vaccination pre-enrollment | No | 160 | 0.000 | - |  |
|  | Yes | 108 | 0.441 | (0.069, 0.813) | 0.0022 |
| HCT-CI | 0 | 60 | 0.000 | - |  |
|  | ≥1 | 208 | -0.358 | (-0.729, 0.012) | 0.0127 |
| B cell and T cell counts measured pre-enrollment | No | 73 | .000 | - |  |
|  | Yes | 195 | -0.381 | (-0.766, 0.004) | 0.0109 |
| Log_10_ B cell count in measured patients^c^ | - | 268 | 0.265 | (0.009, 0.520) | 0.0076 |
| Log_10_ T cell count in measured patients^c^ | - | 268 | 0.345 | (-0.036, 0.726) | 0.0197 |
| N indicates number; CI indicates confidence interval. Estimates >0 indicate a positive association and <0 indicate a negative association with the outcome of anti-Spike IgG level on a log_10_ scale. A stepwise variable selection procedure was used to determine covariates to include in the model, with a p-value < 0.05 as the criterion for inclusion. Cellular therapy and vaccine initation timing were included in all models. The displayed variables were the only variables retained in the adjusted models. Potential interactions were evaluated between covariates. Patients who received Evusheld before the visit’s sample collection were excluded from the analysis. ^a^Covariates considered in models included baseline and time dependent variables from Tables 1 and 2. Time-dependent variables were considered in models at baseline as well as at the time point preceding each endpoint.  ^b^Adjusted response rate estimates and confidence intervals were constructed for each cellular therapy and vaccine timing subgroup and compared between timing groups within cell therapies. No significant differences were observed.  ^c^Median B cell count was 84.19 cells/uL; median T cell count was 272.35 cells/uL. | | | | | |

| **Table S8.** Adjusted logistic and linear regression models of the association of baseline and time-dependent variables with SARS-CoV-2-specific T cell response at the post-V2 time point. | | | | | |
| --- | --- | --- | --- | --- | --- |
| **Variable^a^** | **Category** | **N** | **Odds Ratio or Estimate** | **99% CI** | **p-value** |
| **Logistic regression model of qualitative T cell response (T-Detect assay positive)** | | | | | |
| Cellular therapy and vaccine timing categories^b^ | AlloHCT, < 4 Months | 15 | 1.000 | - | 0.1795 (5df) |
|  | AlloHCT, 4 - 12 Months | 32 | 0.927 | (0.151, 5.706) | 0.9139 |
|  | AutoHCT, < 4 Months | 21 | 6.062 | (0.283, 130.01) | 0.1300 |
|  | AutoHCT, 4 - 12 Months | 26 | 2.239 | (0.243, 20.656) | 0.3500 |
|  | CAR-T, < 4 Months | 13 | 0.516 | (0.041, 6.515) | 0.5016 |
|  | CAR-T, 4 - 12 Months | 17 | 0.345 | (0.032, 3.717) | 0.2487 |
| Recipient SARS-CoV-2 vaccination pre-enrollment | No | 85 | 1.000 | - |  |
|  | Yes | 39 | 5.292 | (0.743, 37.721) | 0.0289 |
| **Linear regression model of SARS-CoV-2-specific T cell breadth (x10^3^)** | | | | | |
| Vaccine timing cohort^b^ | AlloHCT, < 4 Months | 18 | 0.000 | - | 0.4637 (5df) |
|  | AlloHCT, 4 - 12 Months | 42 | -0.089 | (-0.376, 0.198) | 0.4248 |
|  | AutoHCT, < 4 Months | 24 | 0.001 | (-0.317, 0.319) | 0.9949 |
|  | AutoHCT, 4 - 12 Months | 29 | -0.111 | (-0.422, 0.199) | 0.3564 |
|  | CAR-T, < 4 Months | 17 | -0.199 | (-0.560, 0.162) | 0.1547 |
|  | CAR-T, 4 - 12 Months | 19 | -0.203 | (-0.548, 0.141) | 0.1287 |
| Recipient SARS-CoV-2 infection pre-enrollment | No | 129 | 0.000 | - |  |
|  | Yes | 20 | 0.507 | (0.255, 0.758) | < 0.0001 |
| HCT-CI | 0 | 33 | 0.000 | - |  |
|  | ≥1 | 115 | -0.178 | (-0.390, 0.033) | 0.0298 |
| **Linear regression model of SARS-CoV-2-specific T cell depth (x10^3^)** | | | | | |
| Cellular therapy and vaccine timing categories^b^ | AlloHCT, < 4 Months | 18 | 0.000 | - | 0.0221 (5df) |
|  | AlloHCT, 4 - 12 Months | 42 | -26.988 | (-82.987, 29.011) | 0.2145 |
|  | AutoHCT, < 4 Months | 24 | 19.820 | (-42.178, 81.818) | 0.4102 |
|  | AutoHCT, 4 - 12 Months | 29 | -11.966 | (-72.527, 48.594) | 0.6108 |
|  | CAR-T, < 4 Months | 17 | -12.845 | (-83.249, 57.560) | 0.6384 |
|  | CAR-T, 4 - 12 Months | 19 | -60.875 | (-128.092, 6.343) | 0.0197 |
| Recipient SARS-CoV-2 infection pre-enrollment | No | 129 | 0.000 | - |  |
|  | Yes | 20 | 75.513 | (26.432, 124.594) | < 0.0001 |
| HCT-CI | 0 | 33 | 0.000 | - |  |
|  | ≥1 | 115 | -44.252 | (-85.460, -3.045) | 0.0057 |
| N indicates number; CI indicates confidence interval.  A stepwise variable selection procedure was used to determine covariates to include in the model, with a p-value < 0.05 as the criterion for inclusion. Cellular therapy and vaccine initation timing were included in all models. The displayed variables were the only variables retained in the adjusted models. Potential interactions were evaluated between covariates. ^a^Covariates considered in models included baseline and time dependent variables from Tables 1 and 2. Time-dependent variables were considered in models at baseline as well as at the time point preceding each endpoint.  ^b^Adjusted response rate estimates and confidence intervals were constructed for each cellular therapy and vaccine timing subgroup and compared between timing groups within a cell therapy cohort (e.g., among allogeneic HCT only, autologous HCT only, and CAR-T cell therapy only). No significant differences were observed. | | | | | |

| **Table S9.** Comparison of SARS-CoV-2 antibody and T Cell metrics among participants with (cases) and without (controls) subsequent SARS-CoV-2 infection. | | | | | | |
| --- | --- | --- | --- | --- | --- | --- |
| **Outcome Variable** | **Group** | **N** | **Median** | **Interquartile Range** | **Range** | **p-value** |
| SARS-CoV-2 infections between post-vaccination 2 time point and end-of-study | | | | | | |
| Anti-spike Ab (U/mL) | Cases | 38 | 266.0 | (31.8 - 2501) | (0.2 – 2501) | 0.053 |
|  | Controls | 348 | 1967.5 | (125 - 2501) | (0.2 – 2501) |  |
| Neutralizing D614G Ab Titers^a^ | Cases | 16 | 8449.4 | (302.0 – 16577.3) | (39.9 – 81092.8) | 0.987 |
|  | Controls | 122 | 3670.6 | (419.4 – 23556.5) | (39.9 – 709237.0) |  |
| Neutralizing Delta Ab Titers^a^ | Cases | 16 | 3368.0 | (214.6 – 17403.9) | (39.9 – 76206.2) | 0.752 |
|  | Controls | 122 | 1826.0 | (146.8 – 11454.0) | (39.9 – 628946.4) |  |
| Neutralizing Omicron Ab Titers^a^ | Cases | 16 | 39.9 | (39.9 – 91.6) | (39.9 – 608.2) | 0.138 |
|  | Controls | 122 | 62.3 | (39.9 – 434.93) | (39.9 – 167098.5) |  |
| Absolute Lymphocyte Count | Cases | 33 | 1300.0 | (670.0, 1700.0) | (200.0, 3700.0) | 0.170 |
|  | Controls | 330 | 1000.0 | (600.0, 1500.0) | (0.0, 12000.0) |  |
| SARS-CoV-2 T Cell Breadth, 10^6^ units^a^ | Cases | 19 | 174.2 | (103.7, 513.9) | (36.8, 2557.0) | 0.146 |
|  | Controls | 130 | 253.7 | (146.7, 506.2) | (0.0, 3068.2) |  |
| SARS-CoV-2 T Cell Depth, 10^6^ units^a^ | Cases | 19 | 13.5 | (-9,7, 95.9) | (-14.2, 182.9) | 0.643 |
|  | Controls | 130 | 38.0 | (7.4, 68.0) | (-19.4, 518.4) |  |
| Anti-S IgG >2,500 U/mL | Cases | 13/176 (3.4%) | -- | -- | -- | 0.159 |
|  | Controls | 25/210 (6.5%) | -- | -- | -- |  |
| T-Detect Positive | Cases | 9/98 (9%) | -- | -- | -- | 0.128 |
|  | Controls | 10/51 (20%) | -- | -- | -- |  |
| SARS-CoV-2 infections between end-of-study and up to 6 months thereafter^b^ | | | | | | |
| Anti-spike Ab (U/mL) | Cases | 10 | 1734.5 | (703, 2501) | (0.7, 2501) | 0.056 |
|  | Controls | 258 | 5952.5 | (1020, 2501) | (0.7, 2501) |  |
| Anti-S IgG >2,500 U/mL | Cases | 4/274 (1.5%) | -- | -- | -- | 0.098 |
|  | Controls | 6/94 (2.2%) | -- | -- | -- |  |
| Comparisons are based on Wilcoxon rank sum tests or Fisher’s exact test as appropriate. Patients who received tixagevimab-cilgavimab (Evusheld) within the prior six months were excluded from antibody analyses.  ^a^Neutralizing antibody titers and SARS-CoV-2 specific T cells were measured in a subgroup of participants.  ^b^Only results for anti-S IgG are shown, as this was available for all participants and the number of SARS-CoV-2 infections was too low in the subgroup with other immune metrics. | | | | | | |

| **Table S10.** New graft-versus-host disease (GVHD) events and grade 3 or higher adverse events possibly related to SARS-CoV-2 mRNA vaccination reported at each time point. | | | | | | | | | | | | | | | | | | | | |
| --- | --- | --- | --- | --- | --- | --- | --- | --- | --- | --- | --- | --- | --- | --- | --- | --- | --- | --- | --- | --- |
|  | ***AlloHCT*** | | | | | ***AutoHCT*** | | | | | ***CAR-T*** | | | | | ***Overall*** | | | | |
|  | ***V2*** | ***V3*** | ***V3B*** | ***V4*** | ***V5*** | ***V2*** | ***V3*** | ***V3B*** | ***V4*** | ***V5*** | ***V2*** | ***V3*** | ***V3B*** | ***V4*** | ***V5*** | ***V2*** | ***V3*** | ***V3B*** | ***V4*** | ***V5*** |
| ***Total*** | 225 | 213 | 49 | 157 | 193 | 156 | 152 | 29 | 117 | 137 | 60 | 57 | 10 | 46 | 55 | 441 | 422 | 88 | 320 | 385 |
| ***New-onset GVHD*** |  |  |  |  |  |  |  |  |  |  |  |  |  |  |  |  |  |  |  |  |
| ***aGVHD skin stage*** |  |  |  |  |  |  |  |  |  |  |  |  |  |  |  |  |  |  |  |  |
| *1* | 3 | 1 | 1 | 3 | 3 | - | - | - | - | - | - | - | - | - | - | 3 | 1 | 1 | 3 | 3 |
| *2* | - | 2 | 1 | 1 | 3 | - | - | - | - | - | - | - | - | - | - | - | 2 | 1 | 1 | 3 |
| *3* | 2 | - | - | 2 | - | - | - | - | - | - | - | - | - | - | - | 2 | - | - | 2 | - |
| ***Upper GI abnormalities*** | 1 | 1 | - | 2 | 2 | - | - | - | - | - | - | - | - | - | - | 1 | 1 | - | 2 | 2 |
| ***aGVHD lower GI stage*** |  |  |  |  |  |  |  |  |  |  |  |  |  |  |  |  |  |  |  |  |
| *1* | - | 1 | - | 1 | - | - | - | - | - | - | - | - | - | - | - | - | 1 | - | 1 | - |
| *3* | 1 | - | - | 1 | - | - | - | - | - | - | - | - | - | - | - | 1 | - | - | 1 | - |
| *4* | - | 1 | - | - | - | - | - | - | - | - | - | - | - | - | - | - | 1 | - | - | - |
| ***aGVHD liver stage*** |  |  |  |  |  |  |  |  |  |  |  |  |  |  |  |  |  |  |  |  |
| *1* | 1 | - | - | - | - | - | - | - | - | - | - | - | - | - | - | 1 | - | - | - | - |
| *2* | - | - | - | - | 1 | - | - | - | - | - | - | - | - | - | - | - | - | - | - | 1 |
| ***cGVHD onset type*** |  |  |  |  |  |  |  |  |  |  |  |  |  |  |  |  |  |  |  |  |
| *Progressive* | 1 | 1 | 1 | 3 | 3 | - | - | - | - | - | - | - | - | - | - | 1 | 1 | 1 | 3 | 3 |
| *Interrupted* | 3 | 6 | 1 | 2 | 10 | - | - | - | - | - | - | - | - | - | - | 3 | 6 | 1 | 2 | 10 |
| *De novo* | 6 | 6 | - | 2 | 11 | - | - | - | - | - | - | - | - | - | - | 6 | 6 | - | 2 | 11 |
| ***cGVHD maximum grade*** |  |  |  |  |  |  |  |  |  |  |  |  |  |  |  |  |  |  |  |  |
| *Mild* | 6 | 7 | 2 | 3 | 16 | - | - | - | - | - | - | - | - | - | - | 6 | 7 | 2 | 3 | 16 |
| *Moderate* | 4 | 6 | - | 4 | 6 | - | - | - | - | - | - | - | - | - | - | 4 | 6 | - | 4 | 6 |
| *Severe* | - | - | - | - | 2 | - | - | - | - | - | - | - | - | - | - | - | - | - | - | 2 |
| ***cGVHD limited or extensive*** |  |  |  |  |  |  |  |  |  |  |  |  |  |  |  |  |  |  |  |  |
| *Limited* | 7 | 6 | - | 3 | 13 | - | - | - | - | - | - | - | - | - | - | 7 | 6 | - | 3 | 13 |
| *Extensive* | 3 | 7 | 2 | 4 | 11 | - | - | - | - | - | - | - | - | - | - | 3 | 7 | 2 | 4 | 11 |
| ***Adverse events ≥ Grade 3*** |  |  |  |  |  |  |  |  |  |  |  |  |  |  |  |  |  |  |  |  |
| *Fever* | - | 1 | 1 | - | - | - | - | - | - | - | - | - | - | - | - | - | 1 | 1 | - | - |
| *Fatigue* | 1 | 2 | - | - | - | - | 1 | - | - | - | - | - | - | - | - | 1 | 3 | - | - | - |
| *Allergic reaction* | - | - | - | 1 | 2 | - | 1 | - | 1 | - | - | - | - | - | - | - | 1 | - | 2 | 2 |
| *Nausea* | - | 1 | 1 | - | 2 | 1 | - | - | - | - | - | - | - | - | - | 1 | 1 | 1 | - | 2 |
| *Vomiting* | 1 | - | - | - | 2 | - | - | - | - | - | - | - | - | - | - | 1 | - | - | - | 2 |
| *Diarrhea* | - | 2 | - | - | 1 | - | 1 | - | - | 1 | - | - | - | - | - | - | 3 | - | - | 2 |
| *Cystitis noninfective* | - | - | - | - | 1 | - | - | 1 | - | - | - | - | - | - | - | - | - | 1 | - | 1 |
| *Acute kidney injury* | - | - | - | 1 | 1 | - | - | - | - | - | - | - | - | - | - | - | - | - | 1 | 1 |
| *Chronic kidney disease* | - | - | - | 1 | 1 | - | - | - | - | 1 | - | - | - | - | - | - | - | - | 1 | 2 |
| *Dialysis* | - | 1 | - | 2 | 1 | - | - | - | - | - | - | - | - | - | - | - | 1 | - | 2 | 1 |
| *Hypotension* | - | 1 | - | 1 | 2 | - | - | - | - | - | - | - | - | - | - | - | 1 | - | 1 | 2 |
| *Hypertension* | 2 | 3 | - | - | - | - | 1 | - | - | 1 | 1 | 1 | - | - | 2 | 3 | 5 | - | - | 3 |
| *Myocardial infarction* | - | - | - | - | - | 1 | - | - | - | - | - | - | - | - | - | 1 | - | - | - | - |
| *Pericardial effusion* | 1 | - | - | - | 1 | 1 | - | - | - | - | - | - | - | - | - | 2 | - | - | - | 1 |
| *Restrictive cardiomyopathy* | - | - | - | - | - | - | 1 | - | - | - | - | - | - | - | - | - | 1 | - | - | - |
| *Somnolence* | - | - | - | 1 | - | - | 1 | - | 1 | - | - | - | - | - | - | - | 1 | - | 2 | - |
| *RPLS/PRES* | - | - | - | - | - | - | 1 | - | - | - | - | - | - | - | - | - | 1 | - | - | - |
| *TTP / TMA* | - | - | - | 1 | - | - | - | - | - | 1 | - | - | - | - | 1 | - | - | - | 1 | 2 |
| *Thromboembolic event* | - | 1 | - | - | - | - | - | - | 1 | 1 | - | - | - | - | - | - | 1 | - | 1 | 1 |
| *Avascular necrosis* | - | - | - | - | - | - | - | - | - | 1 | - | - | - | - | - | - | - | - | - | 1 |
| *Arthralgia (joint pain)* | - | - | 1 | - | - | - | - | - | - | - | - | - | - | - | - | - | - | 1 | - | - |
| *Myalgia (muscle pain)* | - | 1 | 1 | - | 1 | - | - | - | 1 | - | - | - | - | - | - | - | 1 | 1 | 1 | 1 |
| *Hypoxia* | - | - | - | 1 | 3 | - | - | - | - | - | - | - | - | - | - | - | - | - | 1 | 3 |
| *Dyspnea* | - | - | - | 2 | 1 | - | - | - | - | 1 | - | - | - | - | - | - | - | - | 2 | 2 |
| *Hyperglycemia* | - | - | - | 2 | 2 | - | - | - | - | 1 | - | - | - | - | 1 | - | - | - | 2 | 4 |
| *Hepatitis* | 7 | 3 | - | - | - | - | 1 | - | - | - | - | - | - | - | - | 7 | 4 | - | - | - |
| V2, V3, V4, and V5 indicate the post-V1, post-V2, post-V3, and end-of-study time points, respectively. V3B was an additional post-V2 time point obtained in a subset of participants who had prolonged durations in between vaccines due to changes in vaccine recommendations when additional doses were recommended. Adverse events (according to NCI CTCAE Version 5.0) are shown for those documented prior to the indicated time point. Participants may have the same adverse event in multiple visits. | | | | | | | | | | | | | | | | | | | | |

| Table S11. Antibodies used for flow cytometry. | | |
| --- | --- | --- |
| Specificity | **Clone** | **Fluorophore** |
| CD69 | FN50 | BV750 |
| amine-reactive | n/a | LIVE/DEAD Blue |
| CD45RA | 5H9 | BUV395 |
| CD14 | MφP9 | BUV563 |
| CD45RO | UCHL1 | BUV805 |
| CD8 | 3B5 | Qdot800 |
| CD19 | SJ25-C1 | Qdot 605 |
| CD11c | B-ly6 | BUV661 |
| HLA-DR | L243 | BV570 |
| CD45 | 2D1 | PerCP |
| CD4 | SK3 | APC Fire 810 |
| CD3 | OKT3 | BV510 |

### REFERENCES

1.	Huang Y, Borisov O, Kee JJ, et al. Calibration of two validated SARS-CoV-2 pseudovirus neutralization assays for COVID-19 vaccine evaluation. Sci Rep [Internet] 2021;11(1):1–13. Available from: https://doi.org/10.1038/s41598-021-03154-6

2.	Van Gassen S, Callebaut B, Van Helden MJ, et al. FlowSOM: Using self-organizing maps for visualization and interpretation of cytometry data. Cytom Part A 2015;87(7):636–45.

3.	Amir E ad D, Lee B, Badoual P, et al. Development of a comprehensive antibody staining database using a standardized analytics pipeline. Front Immunol 2019;10(JUN):1315.

4.	Finak G, Langweiler M, Jaimes M, et al. Standardizing Flow Cytometry Immunophenotyping Analysis from the Human ImmunoPhenotyping Consortium. Sci Rep [Internet] 2016 [cited 2022 Nov 21];6. Available from: https://pubmed.ncbi.nlm.nih.gov/26861911/

5.	Maecker HT, McCoy JP, Nussenblatt R. Standardizing immunophenotyping for the Human Immunology Project. Nat Rev Immunol 2012 123 [Internet] 2012 [cited 2022 Nov 21];12(3):191–200. Available from: https://www.nature.com/articles/nri3158
